# Consensus guidelines for eligibility assessment of pathogenic variants to antisense oligonucleotide treatments

**DOI:** 10.1101/2024.09.27.24314122

**Authors:** David Cheerie, Margaret Meserve, Danique Beijer, Charu Kaiwar, Logan Newton, Ana Lisa Taylor Tavares, Aubrie Soucy Verran, Emma Sherrill, Stefanie Leonard, Stephan J. Sanders, Emily Blake, Nour Elkhateeb, Aastha Gandhi, Nicole S. Y. Liang, Jack T. Morgan, Anna Verwillow, Jan Verheijen, Andrew Giles, Sean Williams, Maya Chopra, Laura Croft, Hormos Salimi Dafsari, Alice E. Davidson, Jennifer Friedman, Anne Gregor, Bushra Haque, Rosan Lechner, Kylie-Ann Montgomery, Mina Ryten, Emil Schober, Gabriele Siegel, Patricia Sullivan, Bianca Zardetto, Timothy Yu, Matthis Synofzik, Annemieke Aartsma-Rus, Gregory Costain, Marlen C. Lauffer, the N=1 Collaborative

## Abstract

Of the around 7,000 known rare diseases worldwide, disease-modifying treatments are available for fewer than 5%, leaving millions of individuals without specialized therapeutic strategies. In recent years, antisense oligonucleotides (ASOs) have shown promise as individualized genetic interventions for rare genetic diseases. However, there is currently no consensus on which disease-causing DNA variants are suitable candidates for this type of genetic therapy. The Patient Identification Working Group of the N=1 Collaborative (N1C), alongside an international group of volunteer assessors, has developed and piloted consensus guidelines for assessing the eligibility of pathogenic variants towards ASO treatments. We herein present the N1C VARIANT (**V**ariant **A**ssessments towa**r**ds El**i**gibility for **An**tisense Oligonucleotide **T**reatment) guidelines, including the guiding scientific principles and our approach to consensus building. Pathogenic, disease-causing variants can be assessed for the three currently best-established ASO treatment approaches: splice correction, exon skipping, and downregulation of RNA transcripts. A genetic variant is classified as either “eligible”, “likely eligible”, “unlikely eligible”, or “not eligible” in relation to the different approaches, or “unable to assess”. We also review key considerations for assessment for upregulation of transcripts from the wildtype allele, an emerging ASO therapeutic strategy. We provide additional tools and training material to enable clinicians and researchers to use these guidelines for their eligibility assessments. With this initial edition of our N1C VARIANT guidelines, we provide the rare genetic disease community with guidance on how to identify suitable candidates for variant-specific ASO-based therapies and the possibility of integrating such assessments into routine clinical practice.

## Introduction

There are about 7,000 different rare diseases known to date, with disease-modifying treatments approved for about 5% of them.^1, 2^ A rare disease is defined as a condition that affects less than 200,000 people in the US, or less than 1 in 2,000 individuals within Europe and Canada.^1^ It is estimated that 6% of the world’s population lives with a rare disease.^3^ The majority of rare diseases are thought to be genetic in origin, and with the massive improvements made in genetic diagnostics in the last decades, we can now diagnose up to 50% of individuals who suffer from a rare disease.^4^ As more individuals receive a molecular genetic diagnosis, the need to develop targeted treatments is increasingly urgent. Yet, with many of these rare diseases only affecting a handful of individuals across the globe, the usual drug development route is not a viable pathway in most cases and more bespoke therapeutic strategies are necessary.^5^

Antisense oligonucleotides (ASOs) are one promising form of genetic therapy. Over 20 different oligonucleotide therapies for general applications have been approved by either the Food and Drug Administration (FDA), European Medicines Agency (EMA), the UK’s Medicines & Healthcare products Regulatory Agency (MHRA) and/or the Japanese Ministry of Health, Labour and Welfare^6^ and these drugs have been administered and well-studied in thousands of patients worldwide. Systemic delivery is possible (for instance, via subcutaneous or intravenous injection) but localized or targeted delivery is also feasible for a growing number of target organs (brain and spinal cord via intrathecal injection, eye via intraocular injection, liver, and muscle via GalNAc and transferrin receptor targeting respectively), allowing relatively low doses to be administered with potentially high treatment effects.^7^ Due to the finite half-life of ASOs, treatment needs to be administered repeatedly (often every 1-4 months), but this also allows treatment regimen and dosing to be tailored for each individual where helpful, optimizing to individual benefit and side effects.

Since 2018, multiple groups and organizations have developed ASO treatments for individuals and small groups of patients, either targeted to their specific variant, a single nucleotide polymorphism, or the disease gene in general.^8-10^ These developments have given hope to the rare disease community that individualized, disease-modifying therapies may be a realistic option for additional patients in the near future. More than 10 individuals have received individualized ASO therapies, and more are under development (personal communication).

ASOs are versatile in their usage, as they can be employed to: (i) downregulate transcripts in the case of toxic gain-of-function (GoF) and dominant negative variants, (ii) restore the reading frame in case of truncating variants leading to a loss-of-function (LoF) effect, (iii) correct aberrant splicing, and (iv) increase protein expression of the wildtype allele in disorders associated with haploinsufficiency (see Suppl. File 1).^11^ Yet, not all genetic variants can be targeted with ASOs and even the ones that can be targeted can be distinguished into more eligible (stronger) and less eligible (weaker) candidates. It is thus important to systematically assess every case for its eligibility for ASO treatment to identify the individuals most likely to benefit from such therapies.

The N=1 Collaborative (N1C) (https://www.n1collaborative.org/) is a global initiative to standardize ultra-rare “n-of-1/few” therapy development and deliver it safely and equitably to individuals with rare diseases. The N1C Patient Identification Working Group (PIWG) is focused on three key areas: (i) identifying suitable genetic variants for ASO development, (ii) determining diseases that are prime candidates for genetic therapy, and (iii) selecting patients who are suitable for individualized genetic therapy development. Additionally, the group aims to provide structured informational support to clinicians, researchers, and patient organizations on identifying individuals most likely to benefit from genetic therapies, understanding the necessary disease information to inform therapeutic decisions, and providing guidance on patient communication throughout the development process. The PIWG has developed criteria and established a consensus on how to assess diagnostic DNA variants for amenability to ASO therapies. The guidelines are based on two recently published frameworks from members within the group^9,12^, and this work has now been extended beyond the initial frameworks to include criteria for many types of genetic pathological mechanisms causing a monogenic disorder.

Here, we describe the development of the consensus guidelines - named N1C VARIANT (**V**ariant **A**ssessments towa**r**ds El**i**gibility for **An**tisense Oligonucleotide **T**reatment) guidelines -, present the first version of the guidelines for use by the wider community, and provide training material such as example assessments and training videos. We further introduce the “N1C Variant Eligibility Calculator” that aids with the assessments.

## Methods

### Overview of Guideline Development

The development of the consensus guidelines (N1C VARIANT Guidelines) was a multisite effort with input from pre-clinical and clinical researchers, and genetics healthcare providers. The guidelines were developed through alternating rounds of revisions and piloting, leading to the final version 1.0 (Suppl. File 1).

#### Version 0.1

Development began with a PIWG internal assessment of sample variants. A single assessor from four participating sites [the Dutch Center for RNA Therapeutics (DCRT), The Netherlands; Hospital for Sick Children (SickKids), Toronto, Canada; the Hertie-Institute for Clinical Brain Research, Tübingen, Germany; Boston Children’s Hospital, USA] independently assessed 30 selected variants (previously assessed at the DCRT). The assessment approaches and outcomes from each site were compared, debated by the PIWG, and distilled into an outline of the guidelines.

This outline of the guidelines for ASO eligibility proposed the purpose, content, format, and definitions of classifications. The outline was shared with the PIWG membership for input and revised based on their feedback. This outline was used to draft the first version of the consensus guidelines (version 0.1). This first version was only applicable to LoF variants in genes causing autosomal recessive and X-linked recessive disorders, and only assessed variants for exon skipping and splice correcting ASOs. This draft was shared with the PIWG, where feedback was collected and applied. Following revisions by the PIWG, the revised draft was shared with a group of external volunteers (n=5), who reviewed the guidelines, provided feedback, and assessed a test set of three variants (Suppl. File 2).

#### Version 0.2

Feedback and assessment results from the external volunteers were collected as written responses and used for further revision (version 0.2). We paid attention not only to the feedback on the guidelines but also to how the test variants were assessed and whether the reasoning for the assessments was in alignment with our guidelines. When assessors did not come to the correct conclusion we rephrased and adjusted the guidelines to aid with the assessments.

Version 0.2 was once again shared with the PIWG for edits and feedback before being distributed to a larger group of external volunteer assessors (n=14) for a second round of piloting on a set of twelve test variants (Suppl. File 2). Once again, feedback and assessment reports were collected and used to revise the guidelines.

#### Version 0.3

In version 0.3, the guidelines were expanded to include assessment of eligibility towards ASO/siRNA-mediated transcript knockdown for GoF and dominant negative variants, and towards upregulation from the WT allele (e.g., targeted augmentation of nuclear gene output [TANGO]^13^) as well as all known types of inheritance patterns. This revised guideline was shared with external volunteers (n=19) for a third round of piloting with 15 test variants (Suppl. File 2). Assessment results and feedback were collected as written responses, in addition to the explanations of their assessments, and together used by the PIWG to refine the guidelines. This final version (version 1.0) was shared with all co-authors for final feedback before submission.

### Test Variant Curation

All test variants were selected by one member of the PIWG who did not participate in the assessment rounds. Selected variants were sourced from published literature or the ClinVar database (https://www.ncbi.nlm.nih.gov/clinvar/). Two PIWG members independently assessed each variant and determined the feasibility of the variant assessment depending on the publicly available information (including prior success with ASO development, where applicable). Once a variant’s analysis and classification were agreed upon, these members drafted a “correct answer” representing the expected outcome using the guidelines (Suppl. File 3). Answers with explanations were communicated to all volunteer assessors after each assessment round.

### Guidelines Piloting

Volunteer assessors ranged from graduate students (at both the Masters and PhD level) to faculty with different levels of experience in clinical genetics and ASO therapy development. Assessors included trained basic science or translational researchers, genetic counselors, and clinicians. Assessors were recruited via the professional networks connected to the N1C (e.g., departmental colleagues, announcements in the N1C newsletter, and international conferences featuring N1C PIWG members). An overview of where the assessors are currently working and what their role is is given in Table 1.

**Table 1:** Global makeup of volunteer assessors including current institution and role/position.

| Country of Institution | Number of Assessors | Institutions | Role/Position |
| --- | --- | --- | --- |
| Australia | 2 | Children's Cancer Institute | PhD candidate |
|  |  | Queensland University of Technology | Senior Scientist |
| Canada | 4 | The Hospital for Sick Children | Masters research students (3)<br>Genetic Counselor |
| Germany | 3 | Hertie Institute for Clinical Brain Research | Postdoctoral Research Fellow |
|  |  | University of Cologne | Clinician Scientist<br>MD candidate |
| The Netherlands | 2 | Dutch Center for RNA Therapeutics, LUMC | PhD candidate |
|  |  | Erasmus Medical Center | MD/PhD candidate |
| Switzerland | 2 | University Hospital of Bern | Senior Scientist |
|  |  | University of Zurich | Senior Scientist |
| United Kingdom | 4 | University College London | Associate Professor<br>Professor of<br>Neuroscience and<br>Clinical Geneticist<br>Research Associate |
|  |  | Genomics England | Clinical Fellow |
| United States | 8 | Boston Children's Hospital | Genetic Counselor<br>Clinician Scientist |
|  |  | Mayo Clinic | Research Fellow<br>Senior Bioinformatician |
|  |  | Ambry Genetics | Genetic Counselor |
|  |  | Massachusetts General Hospital | Genetic Counselor |
|  |  | Rady Children's Hospital | Clinician Scientist |
|  |  | Children's Hospital Colorado | Clinical Molecular<br>Geneticist |

Once the assessors agreed to participate, they received a welcome email with an introduction to the assessments and initial information on the timeline and tasks.

### Video Example Development

To support the assessor, exemplary assessments of selected test set variants were provided via short videos. The video examples provide step-by-step instructions on assessing variants using the N1C VARIANT guidelines. Videos were designed and recorded on Microsoft PowerPoint. Videos were reviewed by both the PIWG and the volunteer assessors. A subset of videos was first shared with the PIWG, who provided feedback on the content and structure. Videos were then revised before being shared with volunteer assessors during each round of piloting. Feedback on the videos was collected as written responses.

The variants discussed in the video examples were selected by members of the PIWG. The variants were assessed using the N1C VARIANT guidelines. All selected variants were sourced from published literature or the ClinVar database, with some already having a developed ASO. Two members of the PIWG compared analyses and determined the feasibility of the variant assessment approach. Once a variant’s analysis and classification were agreed upon, a step-by-step analysis was recorded – representing a suggested analysis approach to be taken by assessors using the N1C VARIANT guidelines, along with the expected classification of the variant. Videos are available on the N=1 Collaborative YouTube page (https://www.youtube.com/playlist?list=PL1FIwS0tbJHj0-aDMmZ5fUy5d40eiwa8B) and N1C website (https://www.n1collaborative.org/post/n1c-variant-guidelines).

### Development of the N1C Variant Eligibility Calculator

Based on feedback from the assessors and members of the N1C PIWG, an interactive decision tree was developed to facilitate applying the guidelines (the N1C Variant Eligibility Calculator). First, a catalog with questions and answers based on the guidelines was written, including indications of connections between different sections. For the development of an interactive online tool, HTML and Javascript code was written based on the catalog of questions with the help of ChatGPT which provided a skeleton of the code upon request. The tool was deployed on the N1C website (http://eligibilitycalculator.n1collaborative.org/). The full code is available on the N1C’s GitHub page: https://github.com/N1Collaborative/Variant-Eligibility-Calculator. The eligibility calculator was thoroughly tested by several co-authors of this manuscript, including one assessor doing their assessments of the final 15 variants using the tool to see if they came up with the correct answers. Feedback was gathered through a written response and incorporated accordingly.

### Upregulation from the WT allele Table

Our guidelines refer to various resources for the assessment of pathogenic variants towards upregulation from the WT allele (Suppl. File 1), including four landmark papers^13-16^. To aid in the assessment of variants toward upregulation eligibility, a combined file containing the findings from each suggested paper was generated (Suppl. File 4). The data from Mittal *et al*., Lim *et al*., and Felker *et al*., was extracted from the papers’ supplementary files. The data from Liu *et al*. was extracted from the uORF website (http://rnainformatics.org.cn/RiboUORF/) through POST requests. All data from all available genes from each paper was combined into one Excel file. For each gene on the combined file, pLI scores and ClinGen haploinsufficiency (HI) scores were indicated. The pLI score was downloaded from gnomAD v4.0 (https://gnomad.broadinstitute.org/downloads). ClinGen HI scores were downloaded from ClinGen (https://search.clinicalgenome.org/kb/downloads).

## Results

### Purpose of guidelines

We have developed a consensus guideline (N1C VARIANT guideline) for eligibility assessment and prioritization of pathogenic variants to ASO treatments. With these guidelines, assessors can identify genetic variants most likely to benefit from an ASO-based therapy and distinguish these variants from currently less suitable candidates. The full guidelines are available in Supplementary File 1. Updated versions of the guidelines will also be available via the N1C website (https://www.n1collaborative.org/post/n1c-variant-guidelines).

The guidelines take into consideration the genetic diagnosis of the individual, molecular principles, and pathomechanism of the disease and genetic variant. The purpose of the guideline is to provide clinicians, geneticists, and researchers with a framework for analyzing and classifying disease-causing variants for their amenability to ASO-based therapy. With these guidelines, assessors should be able to:

1. Identify variants eligible for assessment and use publicly available databases and resources to assist in the variant assessment process
2. Assess whether a variant is eligible for ASO-mediated splice correction (i.e., correction of mis-splicing)
3. Assess whether a variant is eligible for ASO-mediated exon skipping
4. Assess whether a candidate gene and/or variant is eligible for siRNA or ASO-mediated transcript knockdown
5. Classify variants as either “eligible”, “likely eligible”, “unlikely eligible”, “not eligible” for the aforementioned ASO approaches, or as “unable to assess” using these guidelines. The definition of each classification is dependent on the type of ASO therapy
6. Consider strategies for the upregulation of wildtype alleles in cases of haploinsufficiency

Overall, these guidelines focus on evaluating (likely) pathogenic, disease-causing variants for eligibility towards ASO treatment. For a full assessment of a patient case, disease and individual-specific clinical factors have to also be taken into consideration, but this work is beyond the scope of these guidelines. These guidelines reflect a general way of evaluating variants, but there are many gene, mechanism, and disease-specific considerations and/or exceptions. Throughout the guidelines, relevant exceptions of variant assessments are indicated.

### Guideline Structure

Only a subset of the guidelines will be relevant to evaluating any one specific variant. The guidelines walk readers through a series of steps where they are prompted to verify variant annotations, inheritance patterns, and pathomechanisms. If critical information is unavailable or insufficient, the guidelines prompt readers that this variant is ineligible for further assessment (“unable to assess”). Conversely, if the necessary information is available and known, readers can use the guidelines to identify appropriate or multiple applicable ASO strategies: splice correction, canonical exon skipping, RNA knockdown, or upregulation from the WT allele. Upon identification of a relevant ASO strategy, readers can direct themselves to the relevant section with the help of flowcharts where they assess the variant’s eligibility towards a specific strategy in greater detail. Throughout the guideline, relevant publicly available resources to aid with the assessments are shared.

As part of the guidelines, assessors are encouraged to check whether an ASO has already been developed for a specific variant, exon, or disorder (whether in clinical or preclinical stages). Clear criteria are provided to define what constitutes sufficient evidence for a functional ASO, depending on the strategy. To assist in this search, the guidelines offer resources and recommended search terms for identifying functionally tested ASOs.

At the end of the assessment, assessors can classify a variant’s eligibility towards splice correction, canonical exon skipping, and/or RNA knockdown based on the information gathered using the guidelines. The classification criteria were developed as part of the consenting process and are outlined below. Assessment strategies are provided for upregulation from the WT allele, but classifications towards eligibility are not defined because this area of ASO therapeutics is less established to date. Instead, the guidelines provide context on when upregulation from the WT allele might be used, and considerations for their development. We further provide an easy way to check for multiple approaches at once by combining available resources on upregulation from the WT allele into one simple Excel table (Suppl. File 4).

### Classification terms

The complexity of the assessments necessitated defining new terms for a classification schema. Prior published classification schema assessed variant amenability to ASO therapy using terms such as “probably”, “possibly”, “unlikely” amenable, “exclude from assessment,” and “consider for exon skipping”.^9,12^ However, these schemas focused on only certain types of variants and ASO strategies. To improve on these prior schemas and expand them to multiple ASO strategies, version 1.0 of these guidelines now employs five tiers for all classifications: “eligible”, “likely eligible”, “unlikely eligible”, “not eligible”, and “unable to assess”.

“Eligible” variants are those for which functional evidence supports the effectiveness of an ASO approach. For splice-correcting ASOs, this means that an ASO has already been developed and shown to be effective, either clinically or pre-clinically, for the specific splice-altering variant. In the context of exon skipping, which aims to “skip” the exon containing the pathogenic variant to produce a truncated yet functional protein product, one would search for functional evidence of exon skipping being non-pathogenic. This would include experimentally induced exon skipping events (i.e., CRISPR deletions, ASOs) showing functional evidence at the protein level that residual protein function remains. This also includes naturally occurring exon skipping events (i.e., benign exon skipping events found in healthy populations). A pathogenic variant found within an exon where either experimentally induced or naturally occurring exon skipping occurs would then be classified as eligible for canonical exon skipping ASOs. Similar ideas apply when assessing variants for eligibility towards knockdown. If a gain-of-function or dominant negative variant, both of which can be considered for knockdown, is found on a gene where a non-allele or non-variant specific knockdown approach has been functionally proven, the variant can be classified as eligible towards knockdown ASOs.

“Not eligible” variants are those for which a specific ASO therapeutic approach is not considered possible. This may be due to functional evidence demonstrating the failure of ASO therapies, for example, canonical exon skipping led to a non-functional protein, or molecular criteria that render the variant unsuitable for ASO targeting. Examples of “not eligible” variants include single-exon genes in the context of exon skipping ASOs or genes with tightly regulated dosage in RNA knockdown strategies.

“Likely eligible” variants are those that, based on molecular criteria, could potentially be targeted by an ASO, although no functional evidence is currently available to confirm this. Conversely, “unlikely eligible” variants are those where molecular criteria suggest an ASO is unlikely to be effective, but no functional evidence directly contradicts the potential use of an ASO.

Variants are classified “unable to assess” when they either do not apply to these guidelines, e.g. are of a type that cannot be assessed, or if there is not enough information available that allows for an assessment of the variant, e.g., the inheritance pattern of the variant is unknown or no information on the pathomechanism is available.

### Videos

At the time of publication, 12 videos have been created and shared (Table 2). These videos provide step-by-step guidance for assessors, highlighting key resources and assessment techniques. Each video demonstrates the assessment of a specific variant towards a relevant strategy, with each example leading to a unique outcome. The videos can be found on the N1C YouTube channel (https://www.youtube.com/playlist?list=PL1FIwS0tbJHj0-aDMmZ5fUy5d40eiwa8B) and via the N1C website (https://www.n1collaborative.org/post/n1c-variant-guidelines).

**Table 2:** List of video examples referred to and published with version 1.0 of the N1C VARIANT guidelines. Variants normalized using Mutalyzer (https://mutalyzer.nl/).

| Video | Variant | Gene Symbol | ASO Strategy | Outcome/Classification |
| --- | --- | --- | --- | --- |
| 1 | NM_000350.3:c.2626C>T, p.(Gln876*) | *ABCA4* | Canonical exon skipping | Eligible |
| 2 | NM_016589.4:c.597-1340A>G | *TIMMDC1* | Splice correcting | Eligible |
| 3 | NM_000533.5:c.680dup, p.(Cys228Leufs*5) | *PLP1* | Canonical exon skipping | Not eligible |
| 4 | NM_003793.4:c.213+1G>C | *CTSF* | Splice correcting | Not eligible |
| 5 | NM_000277.3:c.611A>G, p.(Tyr204Cys) | *PAH* | Splice correcting | Unlikely eligible |
| 6 | NM_025152.3:c.815-27T>C | *NUBPL* | Splice correcting | Unlikely eligible |
| 7 | NM_024312.5:c.3503_3504del, p.(Leu1168Glnfs*5) | *GNPTAB* | Canonical exon skipping | Unlikely eligible |
| 8 | NM_001040142.2:c.5645G>A, p.(Arg1882Gln) | *SCN2A* | Knockdown | Eligible |
| 9 | NM_001165963.4:c.3733C>T, p.(Arg1245*)<br>NM_130839.5:c.67C>T, p.(Arg23*) | *SCN1A*<br>*UBE3A* | Upregulation from the WT allele | Eligible |
| 10 | NM_003793.4:c.264del, p.(Cys89Alafs*59) | *CTSF* | Canonical exon skipping | Likely eligible |
| 11 | NM_000170.3:c.538C>T, p.(Gln180*) | *GLDC* | Canonical exon skipping | Not eligible |
| 12 | Multiple variants |  | N/A | Unable to assess |

### Variant Eligibility Calculator

To support the assessment and help assessors focus on going through the sections of the guidelines relevant to their current assessment, the N1C Variant Eligibility Calculator was developed. At the time of publication, the eligibility calculator walks assessors step-by-step through version 1.0 of the N1C VARIANT guidelines. One key feature of the calculator is the inability to skip question prompts. Each step discussed in the guidelines is crucial for in-depth variant assessment and is required for accurate variant classification. Due to the inability to progress without answering the question, users of the calculator are encouraged to further research the gene or variant before proceeding with the assessment. Additionally, the calculator takes into consideration the kind of variants (i.e., missense, stop gain, synonymous) before directing users to relevant sections of the guidelines. Overall, this tool allows users to systematically navigate the guidelines, acting as a “checklist” before proceeding with assessments. The calculator further provides users with the ability to track their assessments and receive a printout of the specific questions answered and what the answers were to understand the overall classification and identify potential mistakes during the assessment process. The calculator is available via the N1C website (http://eligibilitycalculator.n1collaborative.org/) and the N1C’s GitHub page (https://n1collaborative.github.io/Variant-Eligibility-Calculator/).

## Discussion

Here we introduced version 1.0 of the N1C VARIANT (**V**ariant **A**ssessments towa**r**ds El**i**gibility for **An**tisense Oligonucleotide **T**reatment) guidelines for the assessment of pathogenic variants for eligibility towards ASO treatments, alongside the process of their development and consenting. Additionally, we introduce and discuss the development of training materials and tools to aid with variant assessments.

These guidelines represent the first proposed international consensus approach for evaluating the potential eligibility of pathogenic variants causing monogenic disorders toward ASO therapies. With the significant progress and publicity in the last few years regarding the development of individualized genetic therapies^8-10^ there is hope within the rare disease community that ASOs may benefit an increasing number of community members. With our guidelines and the accompanying training materials and tools, we aim to support the rare disease community by providing guidance on which variants are most likely to be eligible for ASO therapies.

As a takeaway from the iterative process of developing these guidelines, we want to emphasize that this is a complex assessment procedure that takes time to learn. Similar to the annotation and classification of pathogenicity for genetic variants with the ACMG guidelines^17^, the assessment of pathogenic variants for their eligibility for ASO treatments takes many different aspects into account.

We propose a new five-tier classification schema for ASO treatment amenability, whereby a variant is classified with respect to specific ASO strategies and can thus receive different labels for different types of ASO treatments. The classifications are “eligible”, “likely eligible”, “unlikely eligible”, “not eligible”, or “unable to assess”. While the “eligible” and “not eligible” definitions are clear, categorizing and classifying variants as “likely eligible” and “unlikely eligible” proves to be more challenging, as the evidence for or against eligibility exists on a spectrum. Future work will aim to refine these categories further, mirroring the efforts to introduce more gradations into the ACMG/AMP variant classification scheme.

We further expect to communicate adjustments in the upcoming years, as technologies advance and knowledge grows regarding which genetic variants are most likely amenable to an ASO treatment. We expect that in some cases variant classifications for a certain ASO approach will change with these adjustments, for example, due to new information on the feasibility of ASO designs for certain types of variants, necessitating reassessments over time. In that regard, we would like to point out that it is important to read the literature on available ASO treatments critically and scrutinize the methodology used and the functional data provided, before considering a variant eligible for ASO treatments.

At this point, there is a limited number of ASOs developed and clinically tested, thus not for all different variant types, pathomechanisms, and inheritance patterns discussed herein are ASOs currently available. That means that no gold-standard evidence for many considerations were available for the establishment of these guidelines, and these guidelines can thus be considered expert opinions. However, the guidelines were based on the collective expertise gained from assessing about 1,500 variants since 2018 at the sites of the PIWG members (BCH, SickKids, DCRT), and the knowledge the PIWG members have on ASO design and development, which involves leading experts in the field of ASO development and tailored ASO therapies.

While efforts were made to test the guidelines with various assessors from different professional backgrounds, the application of these guidelines in diverse healthcare settings might reveal additional needs for adjustments and refinements in the future.

Although the amenability of a specific variant to a certain ASO therapeutic strategy is a fundamental first step, disease- and patient-specific factors are equally important considerations in the development and provision of individualized genetic therapies.^11^ Considerations will also include the reversibility and severity of symptoms. This means that an eligible variant does not necessarily equal an eligible patient. The assessments of disease- and patient-specific factors are outside the scope of this work and will require additional recommendations and guidelines.^18,19^

Ideally, we would like to see the integration of the eligibility assessments into clinical practice, to not only provide individuals suffering from rare diseases with a diagnosis but also with information on possible treatment approaches, where applicable. We believe that with the additional training material and test variants provided, clinical geneticists and specialist human geneticists working in laboratories and diagnostic centers can train themselves to become assessors. In the long term, we envision automation of our guidelines and assessment procedures in the form of tools that take the pathogenic variant as input and deliver an analysis of the best therapeutic strategy for each individual.

We plan to regularly update the guidelines as well as the tools and training material. Updated versions with the version number and date of the update will be provided on the N1C website (https://www.n1collaborative.org/post/n1c-variant-guidelines).

## Data Availability

Data is available in supplementary files 2, 3, and 4 for variant assessments and upregulation from the WT allele approaches. All code was deployed on GitHub under an open-source AGPL license (https://github.com/N1Collaborative/Variant-Eligibility-Calculator).

http://eligibilitycalculator.n1collaborative.org/

https://www.youtube.com/playlist?list=PL1FIwS0tbJHj0-aDMmZ5fUy5d40eiwa8B

https://www.n1collaborative.org/post/n1c-variant-guidelines

## Declaration of interests

Nothing to declare.

## Acknowledgments

We would like to thank further N1C members for their input and help during the development of these guidelines. We especially want to thank Nicole Nolen for all her help and support in making the training material and tools available on the N1C website. We also thank all families and clinicians who have provided us with their genetic diagnoses over the years, which ultimately led to the establishment of these guidelines.

This work was supported by the European Union, project European Rare Disease Research Alliance (ERDERA, # 101156595) (to M.S. and A.A.-R.). D.C. is supported by a SickKids Restracomp Master’s Scholarship and the SickKids Innovators Fund. L.N. is supported by a Canada Graduate Scholarship - Master’s from the Canadian Institutes of Health Research (CIHR). D.C., B.H., and G.C. also acknowledge support from CIHR (PJT186240). D.B. is supported by a Humboldt Research Fellowship and the Hertie-Network of Excellence in Clinical Neuroscience. M.C.L is funded by a Walter Benjamin Fellowship from the DFG. A.E.D, K.-A.M, and M.R. are supported by the UK Platform for Nucleic Acid Therapies (UpNAT). B.Z., A.A.-R., R.L. are supported by a ZonMW PSIDER grant.

## Author contributions

D.C. and M.C.L. of the N1C PIWG conceptualized the N1C VARIANT guidelines. D.C. and M.C.L. drafted the guidelines and the manuscript, conducted the assessment rounds, and evaluated and implemented the feedback. D.C. made the example videos. M.C.L. selected the test variants for assessment and developed and built the variant eligibility calculator.

M.M., D.B., C.K., L.N., A.L.T.T., A.S., E.S., L.N., S.S., M.S., T.Y., A.A.-R., G.C. are members of the N1C PIWG; they reviewed the outlines and drafts of the different rounds of guidelines and edited the manuscript. They further provided feedback on the videos. M.M., D.B., C.K., and L.N., also participated in the variant assessment training and provided feedback and explanations on the assessments. S.L. facilitated the PIWG meetings, set up documents, and helped with putting the work onto the public databases and the N1C website.

E.B., N.E., A.G., N.L., J.M., A.V., J.V., A.G., S.W., M.C., L.C., H.S.D., A.E.D., J.F., A.G., R.L., K.-A.M., M.R., E.S., G.S., P.S. participated as assessors and provided feedback on the guidelines and training material. B.H. made the upregulation from the WT table. L.N. and B.Z. tested the variant eligibility calculator and provided feedback.

Authors were listed as follows: Junior members of the N1C PIWG were named first, followed by assessors who participated in multiple rounds of assessments and provided extensive feedback, all other assessors were named in alphabetical order. Lastly, senior members of the N1C PIWG were named. D.C. and M.C.L. of the N1C PIWG were placed as first and senior authors respectively as they were leading the overall effort.

## Web Resources

Videos available on YouTube: https://www.youtube.com/playlist?list=PL1FIwS0tbJHj0-aDMmZ5fUy5d40eiwa8B

Calculator available on GitHub and N1C website: https://n1collaborative.github.io/Variant-Eligibility-Calculator/ and http://eligibilitycalculator.n1collaborative.org/

Guidelines available on N1C website: https://www.n1collaborative.org/post/n1c-variant-guidelines

## Notes

### Competing Interest Statement

The authors have declared no competing interest.

### Author Declarations

The study used ONLY openly available data from the ClinVar database: https://www.ncbi.nlm.nih.gov/clinvar/

