## Supplemental File 1 for "Consensus guidelines for eligibility assessment of pathogenic variants to antisense oligonucleotide treatments"

***Disclaimer*: This document is the work product of the N=1 Collaborative (the "N1C"). The N1C is not providing legal or regulatory advice for N=1 trials. This document should not be construed as legal or regulatory advice for any particular purpose. These guidelines focus on the evaluation of eligibility of a genetic variant for ASO treatment only. To assess a patient case for ASO amenability, the disease, phenotype, and patient have to also be considered, which is beyond the scope of these guidelines. The information is based on the best available knowledge and practice at the time of publication. The N1C reserves the right to update, modify, or withdraw these practices at any time without prior notice. Users of these documents should exercise their own judgment and discretion in applying these practices to their specific situations. These guidelines reflect a general way of evaluating variants, but there are many exceptions to the rules.**

**The most up-to-date N1C VARIANT Guidelines and all accompanying training material and tools can be found on the N1C website: https://www.n1collaborative.org/post/n1c-variant-guidelines**

### Consensus guidelines for eligibility assessment of pathogenic variants to antisense oligonucleotide treatments: The N1C VARIANT guidelines

**[v1.0 September 2024]**

(written by David Cheerie and Marlen Lauffer on behalf of the N1C Patient Identification Working Group (PIWG))

#### Purpose

The N1C VARIANT (**V**ariant **A**ssessments towa**r**ds El**i**gibility for **An**tisense Oligonucleotide **T**reatment) guidelines provide clinicians, geneticists, and researchers with a framework for analyzing and classifying disease-causing variants for their amenability towards antisense oligonucleotide (ASO) therapies. Specifically, these guidelines are meant to identify genetic variants most likely to benefit from an ASO-based therapy and distinguish these variants from currently less suitable candidates. With these guidelines, assessors should be able to:

1. Identify variants eligible for analysis by these guidelines and utilize publicly available databases and resources to assist in the variant analysis process
2. Identify variants causing aberrant splicing and assess what type of splice-altering variants can be targeted using ASOs
3. Assess whether a variant is eligible for treatment by exon skipping ASOs
4. Assess whether candidate genes and/or variants are eligible for siRNA or ASO-mediated transcript knockdown
5. Classify variants as either “eligible”, “likely eligible”, “unlikely eligible”, “not eligible”, or “unable to assess” towards each of splice correction, exon skipping, or transcript knockdown approaches. The definition of each classified variant is dependent on the type of RNA therapy and is further described in each respective section
6. Consider strategies for upregulation of wildtype alleles in cases of haploinsufficiency

To follow these guidelines, readers should have an understanding of foundational genetic concepts including, but not limited to, splicing, introns and exons, coding versus non-coding, and DNA variant types (nonsense/stop gain, missense, indels, frameshifts, etc.). While these guidelines will remind assessors of key definitions (canonical splice site, cryptic splice site, etc.), these concepts will not be explained in detail. Assessors should also be familiar with standard variant annotations and pathogenicity classification approaches, including preferably the ACMG-AMP guidelines (Richards et al., 2015).

These guidelines focus on the evaluation of eligibility of a pathogenic genetic variant for ASO treatment. To assess a person’s eligibility for ASO treatment, disease- and individual-specific clinical factors have to also be taken into account, which is beyond the scope of these guidelines (Lauffer, van Roon-Mom, Aartsma-Rus, & N = 1 Collaborative, 2024). In some instances, the guidelines do refer to the gene and disease as a necessity for assessment, and this will be pointed out specifically in the respective sections.

The guidelines were prepared to the best of our current knowledge and are subject to change, with new knowledge on the topic being generated continuously. These guidelines reflect a general way of evaluating variants, but there are many exceptions to this. Where necessary, we have mentioned relevant exceptions within the text or as footnotes. This also means a variant's classification can change over time and it might thus be useful to re-assess variants at a later stage. Classifications are made for a variant's eligibility towards a specific ASO strategy. For example, a variant might be classified as "not eligible" for splice correction, while also being classified as "likely eligible" for a transcript knockdown.

We recommend only assessing disease-causing variants classified as likely pathogenic and pathogenic according to the ACMG-AMP guidelines (Richards et al., 2015).

For the assessments, all recommendations on suitable tools, websites, and databases to aid in the process are limited to publicly available resources, but, of course, other licensed resources can also be used at an individual's discretion. Instructions on how to use the recommended tools and websites are beyond the scope of these guidelines. Assessors are encouraged to familiarize themselves with the tools by utilizing the respective tools' "help" pages or corresponding research articles.

#### Structure of guidelines

These guidelines serve as an aid for assessing a given disease-causing variant and are not meant to be read as a whole. Instead, only specific sections need to be read for each assessment. The guideline will first provide the assessor with a background on ASO/siRNA therapies. The guidelines are then divided into 4 steps. Steps 0-2 are necessary to collect information, such as the inheritance pattern or pathomechanism of disease, relevant to each assessment and to decide on the required therapeutic approach. Step 3 is further relevant for each variant assessment and focuses on splicing evaluation, whereas Step 4 provides an overview (Fig. 6) to guide you to relevant sections of the guideline for specific assessments depending on the information gained in Steps 0-3. A quick overview of the structure can be seen in Fig. 4.

Sections A-C can then be read independently and the section matching the variant under assessment can be selected. Within the guidelines, assessors will be provided with the possibility to jump between sections via hyperlinks.

At the end of each section, we have formulated important considerations for each assessment. To support the understanding of the guidelines and their use, we have generated several example assessments and matching training videos as well as a "Variant Eligibility Calculator" that guides through the different steps and sections. The calculator will also aid in identifying the next possible assessment step if a variant is "not eligible" towards one of the strategies.

### Background

ASOs are short, synthetic, single-stranded oligonucleotides that can bind RNA via Watson-Crick base pairing. ASOs can modify protein expression through various mechanisms (Dhuri et al., 2020, Rinaldi & Wood, 2017).

**Splice Modulation**

The binding of ASOs to splice sites or splice-regulatory elements on the pre-mRNA transcript allows for manipulation of the splicing process, which can lead to (canonical) exon skipping or the restoration of wildtype splicing.

ASOs can be used to restore wildtype splicing in individuals whose mechanism of pathogenicity is caused by the activation or creation of cryptic (non-canonical) splice sites, resulting in aberrant splicing of the transcript (Fig. 1A) and, for example, the inclusion of parts of the intron then termed cryptic or pseudoexon. Additionally, canonical exon skipping ASOs can be used to “skip” exons containing the pathogenic variant, to produce a truncated, yet functional, protein product (Fig. 1B) in the case of loss-of-function (LoF) variants. For gain-of-function (GoF) variants, exon skipping can also be applicable, but there is no requirement to generate a functional protein product.

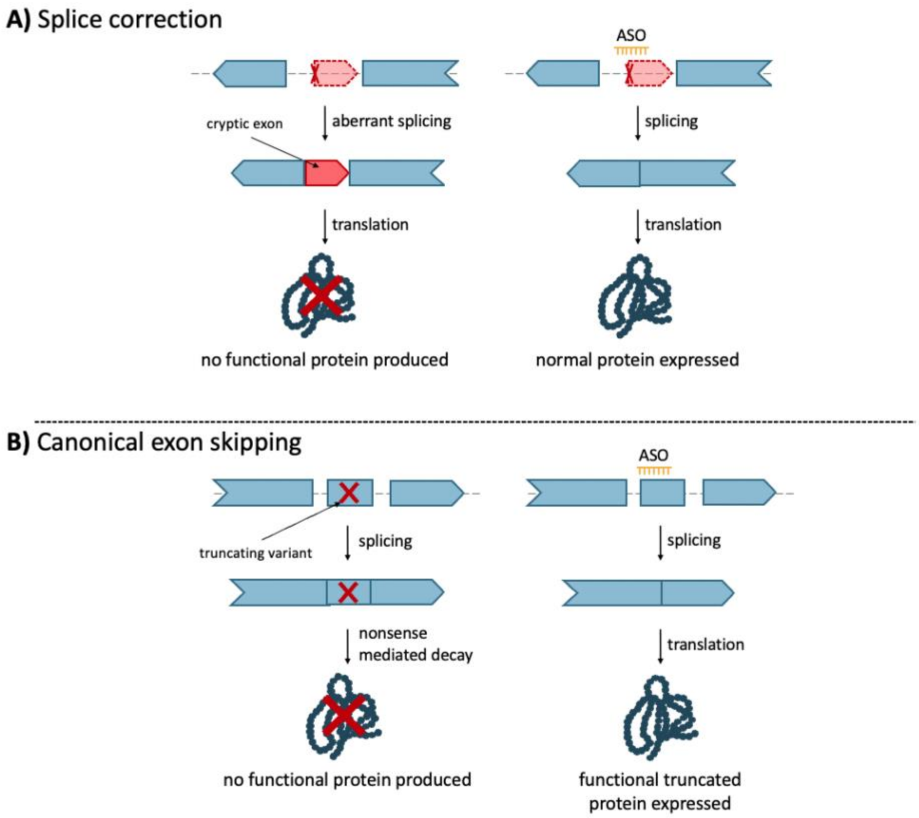

**Figure 1: Splice modulating mechanisms of ASOs for the restoration of a functional gene product.**

**A)** ASOs can be used to skip a cryptic exon caused by deep-intronic pathogenic variants to restore canonical splicing. **B)** ASOs can be used to skip an in-frame canonical exon containing a truncating variant to produce a truncated yet functional gene product.

In rare circumstances, certain variants, like single nucleotide polymorphisms (SNPs) or pathogenic variants, lead to skipping of an exon and subsequently decreased production of protein-coding transcripts. It is possible to develop ASOs that will lead to exon inclusion, however, the development of such efforts is challenging (Singh, Lee, DiDonato, & Singh, 2015).

**Transcript Knockdown**

In addition to splice modulation, oligonucleotides can bind to the target transcript and downregulate (pre-)mRNA expression (i.e., knockdown ASOs). Knockdown can be achieved with gapmer ASOs and small interfering RNAs (siRNAs). Gapmer ASOs trigger RNase H-mediated cleavage (Fig. 2), while siRNAs trigger the endogenous RNA interference pathway. Both mechanisms can be utilized in cases where the pathomechanism is a result of overexpression, toxic GoF, or dominant-negative (DN) effects (Lauffer, van Roon-Mom, Aartsma-Rus, & N = 1 Collaborative, 2024).

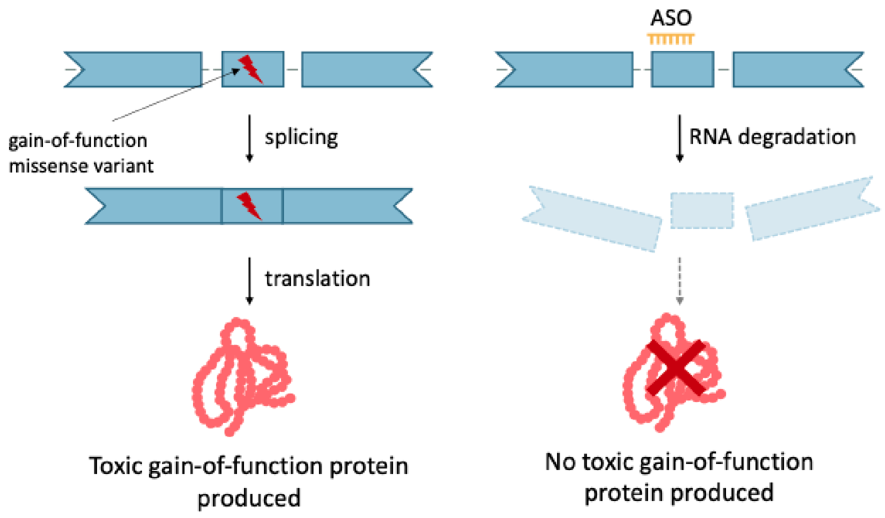

**Figure 2: RNA knockdown using ASOs.**

ASOs can be used to knockdown RNA transcripts which carry variants leading to a toxic GoF protein (shown here), proteins with a DN effect, or overexpressed proteins.

**Increased protein from wildtype transcript**

Furthermore, one can consider upregulation from the WT allele, such as targeted augmentation of nuclear gene output (TANGO), as described in Lim et al., 2020, Mittal et al., 2022, Felker et al., 2023, and Liu et al., 2022 (Fig. 3). For disorders caused by haploinsufficiency, one wildtype allele remains intact and functional. ASOs can be utilized to upregulate the gene product from the wildtype allele with the goal of restoring proper gene and cell function.

One such approach includes the targeting of poison exons. Poison exons are naturally occurring, highly conserved alternatively spliced exons that result in premature termination when included in the transcript. An ASO can be designed to skip poison exons and increase the number of productive/protein-coding transcripts, with the goal of increasing protein levels (Fig. 3A).

Additionally, ASOs can be designed to target naturally occurring antisense transcripts which are non-coding RNAs that can act on one or more corresponding transcripts with diverse roles, including RNA interference and RNA masking (Khorkova et al., 2022). In such cases, targeting antisense transcripts using ASOs can upregulate transcript levels (Fig. 3B).

Lastly, one can target untranslated regions (UTRs) to upregulate or stabilize productive transcripts (Liang et al., 2017; Sasaki et al., 2019). One possible method is the targeting of the upstream open reading frames (uORF). These are alternative reading frames that occur upstream (5') from the canonical reading frame (primary ORF, pORF). These reading frames may code for proteins, but can also downregulate the reading of the canonical reading frame. ASOs targeting the uORF can be used to upregulate transcripts from the canonical reading frame. This can either be done by blocking the uORF or by skipping the exon containing the uORF. A related method is the targeting of the 3' UTR. By using ASOs to interfere with degrading complexes, one can attempt to increase RNA half-life and increase gene product (Fig. 3C).

Overall, the upregulation of wildtype gene products is a possible approach for disorders caused by haploinsufficiency.

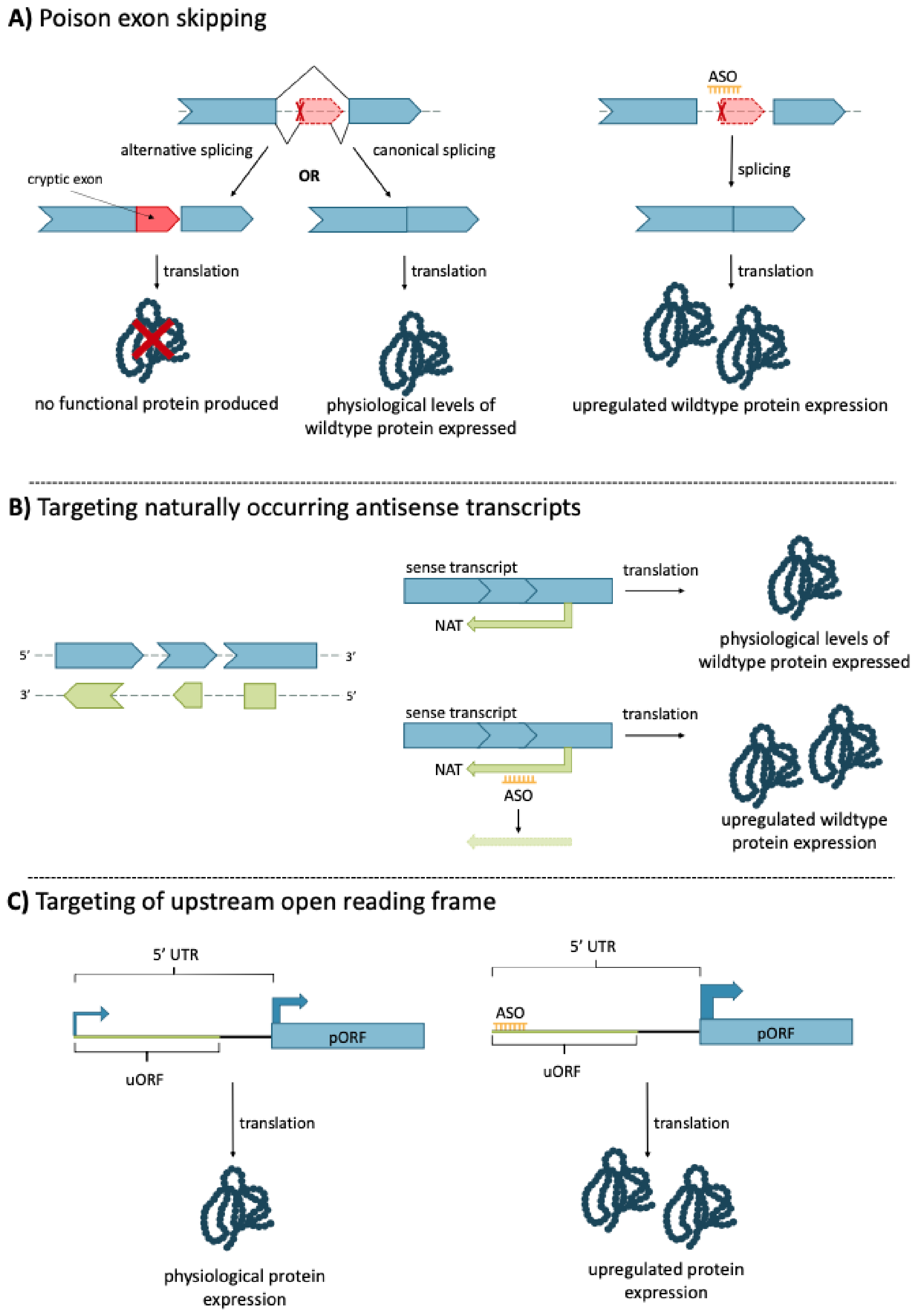

**Figure 3: Upregulation of wildtype allele transcript using ASOs.**

**A)** An ASO can be used to skip a poison exon in a transcript that would usually lead to nonsense-mediated decay, increasing wildtype transcript levels and subsequent gene product. **B)** ASOs can target NATs which negatively impact transcription of the sense transcript. By disrupting NAT transcription, the wildtype transcript can be increased. **C)** Targeting uORFs with an ASO can be used to promote translation of the pORF (primary open reading frame), increasing wildtype gene product.

ASOs offer a potential avenue for the treatment of rare genetic diseases, although this is currently mainly focused on disorders impacting the central nervous system (brain, spinal cord, retina), or liver, due to restricted delivery options. FDA- and EMA-approved ASO therapies include, for example, nusinersen for spinal muscular atrophy (Egli & Manoharan, 2023). The use of ASOs to develop variant-specific therapies has, for example, been demonstrated through milasen, an ASO developed at Boston Children's Hospital, to target a deep intronic *MFSD8* variant resulting in cryptic splicing (Kim et al., 2019).

To better understand the concepts and guidelines shared in this document, it is important to familiarize yourself with the key terms outlined in Table 1.

**Table 1: List of Key Terms**

| **Term** | **Definition** |
|---|---|
| Branchpoint | *cis*-acting intronic motif (specific intronic sequence on the same chromosome) required for pre-mRNA splicing, usually an A (adenosine) located 18-40 nucleotides upstream of the acceptor splice site. |
| Canonical Acceptor Site | RNA sequence recognized by the spliceosome flanking the 3' end of an intron, usually an AU sequence. |
| Canonical Donor Site | RNA sequence recognized by the spliceosome flanking the 5' end of an intron, usually a GU sequence. |
| Canonical Splicing | Splicing involving the use of the canonical acceptor and donor sites (see definitions for canonical acceptor site and canonical donor site). |
| Cryptic Splicing | Cryptic splice sites are naturally occurring splice sites within the genome that are used infrequently. Splicing involving cryptic splice sites, i.e., cryptic splicing, often leads to the incorporation of parts of an intron into the mRNA transcript (cryptic exon or pseudoexon) or the removal of parts of an exon ultimately leading to an early translation stop. Pathogenic variants causing aberrant splicing can for example activate, strengthen, or create a cryptic splice site and thus cause disease. |
| DNA Tandem Repeats | Short lengths of DNA repeated multiple times within a gene, e.g. CAG repeats in PolyQ disorders. |

| | |
|---|---|
| Hypomorphic Allele | Alleles that show partial loss of function. Sometimes referred to as “leaky” alleles because there is some retention of protein function. |
| In-Frame Exon | Exon in which the number of base pairs is divisible by 3. Since each amino acid is encoded in one codon made up of three base pairs, removing an exon that is a multiple of 3 will not disrupt the reading frame. While an exon can be divisible by 3, it does not mean that it starts with the first nucleotide of a codon and ends with the last nucleotide of a codon. The exon boundaries and codon boundaries do not necessarily align. See the “Assessing Exon Position and Frame” section in this document. |
| MANE Select | Matched Annotation from NCBI and EMBL-EBI (MANE): “The MANE Select set consists of one transcript at each protein-coding locus across the genome that is representative of biology at that locus. This set is useful as a universal standard for clinical reporting, as a default for display on browsers and key genomic resources, and as a starting point for comparative or evolutionary genomics. MANE Select transcripts are identified using computational methods complemented by manual review and discussion.’’ (Morales et al., 2022). |
| Naturally Occurring Antisense Transcript | Noncoding antisense transcripts that can act on one or more corresponding transcripts with diverse roles, including RNA interference and RNA masking. |
| Out-of-frame Exon | Exon in which the number of base pairs is not divisible by 3. Please see “in-frame exon” for further explanation on exon frames. |
| Poison Exon | Naturally occurring, highly conserved alternatively spliced exons which result in premature termination when included in the transcript, either as a result of frameshift or the inclusion of a premature stop codon |
| Protein Tandem Repeat Domain | Two or more domains from the same family found in tandem, e.g. spectrin-like repeats in the dystrophin protein |

| | |
|---|---|
| Splicing | A step in the processing of mature mRNA in which introns (non-coding sequences) are removed or “spliced out” of the pre-mRNA transcript, and the remaining exons (coding sequences) are connected to one another forming the mRNA. |
| Splicing Enhancer Site (SE) | RNA sequence motif found in the exon/intron of genes, binds proteins that help recruit splicing machinery to the correct site, directing or enhancing accurate splicing. |
| Splicing Silencer Site (SS) | RNA sequence motif found in the exon/intron of genes, binds proteins that negatively affect the core splicing machinery, inhibiting or silencing the inclusion of an exon into the mRNA. |
| Upstream Open Reading Frame | Alternative reading frames that occur upstream (5’) from the canonical reading frame. |

#### Further Resources

For further resources on the mechanisms and use of ASOs for genetic disorders, please see the below recommendations. Note: these additional resources are not required but are encouraged for those who are not familiar with ASO technology.

Educational videos

Treating Disease at the RNA Level with Oligonucleotides:
https://www.youtube.com/watch?v=nRHypCupg0A

Modifying RNA splicing with Morpholino Oligos by Gene Tools:
https://www.youtube.com/watch?v=qu-Kz0HaLxw&ab_channel=GeneTools

Overview articles

Lauffer et al., *Possibilities and limitations of antisense oligonucleotide therapies for the treatment of monogenic disorders (10.1038/s43856-023-00419-1)* Communications Medicine

Hammond et al., *Delivery of oligonucleotide-based therapeutics: challenges and opportunities* (10.15252/emmm.202013243) EMBO Molecular Medicine

### Variant Assessment

The variant assessment is divided into different steps (please also see flow diagram Fig. 4).
Step 0 - Variant check
Step 1 - Assessment of pattern of inheritance and disease type
Step 2 - Assessment of pathomechanism of genetic variant and haploinsufficiency
Step 3 - Evaluation of splicing effects
Step 4 - Identification of relevant guideline
    Section A - Considerations for Canonical Exon Skipping
    Section B - Considerations for Transcript Knockdown
    Section C - Considerations for upregulation from the wildtype allele

For the assessment of a variant for ASO eligibility, different types of information need to be gathered to decide on suitable ASO strategies and assess a variant using the specific sections (sub-guidelines). To decide on the most suitable guideline for each variant, it is first necessary to check whether the variant can generally be assessed with these guidelines and whether the variant description is correct (Step 0), what the inheritance pattern of the variant and the disorders the gene is associated with are (Step 1), what the pathomechanism of the variant is (Step 2), and whether a variant is influencing splicing (Step 3). With this information at hand, the assessor can decide on possible ASO strategies and read up on the specific sections of the guidelines (Step 4) that will focus on strategies like exon skipping and knockdown approaches.

These guidelines are not meant to be read as a whole, but using the information gathered in Steps 1-3, the flow diagram in Fig. 4, and the detailed diagram in Step 4 (Fig. 6) will guide the assessor to the relevant sections. An exception is Step 3 which not only checks for influence on splicing but in case of effects on splicing already allows the assessor to classify the variant with respect to splice correction ASOs.

We further provide some notable exceptions and special cases as footnotes. These are cases that apply rarely but were added for the assessors to gain a comprehensive understanding of the assessment process.

Variants will be classified as “eligible”, “likely eligible”, “unlikely eligible”, “not eligible”, or “unable to assess” towards a specific approach (Table 2) using these guidelines with the exception of upregulation from the wildtype allele (Section C), where no such classification is possible.

**Table 2: Explanation of Variant Classification Terms**

| Classification | Explanation |
|---|---|
| Eligible | Variants are considered eligible when functional evidence supports the effectiveness of an ASO approach. What type of functional evidence is deemed sufficient depends on the approach and is defined within the different sections below. |

| | |
|---|---|
| Likely eligible | Variants are considered likely eligible variants when the variant could potentially be targeted by an ASO, although no functional evidence is currently available to confirm this. That means a variant meets all criteria relevant for ASO development on paper. |
| Unlikely eligible | Variants are considered unlikely eligible variants when the molecular criteria suggest an ASO is, with our current understanding, unlikely to be effective, but no functional evidence directly contradicts the potential use of an ASO. |
| Not eligible | Variants are considered not eligible when an ASO approach will not work. This can be the case if the genetics do not allow for an ASO correction or an ASO cannot be designed. It could also be that there is evidence that demonstrates an ASO approach will not work. For example, exon skipping leads to a non-functional protein. |
| Unable to assess | All variants that currently cannot be assessed with these guidelines. This can be due to the variant not being applicable to these guidelines or that not enough information is available on a variant that allows for it to be assessed. |

Please note that variants can be applicable and assessed for different approaches and while a variant might be "unlikely eligible" for splice correction, it could for example still be "(likely) eligible" for exon skipping.

The guidelines are also provided with a set of example variant assessments found at the end of this document. Videos accompanying the example assessments walk the assessors through the assessments to aid with training. Assessors can further use the test variants that were assessed during the consenting process for further practice. All test variants can be found in Suppl. File 2 and answer keys are provided in Suppl. File 3.

We have developed the N1C Variant Eligibility Calculator (https://eligibilitycalculator.n1collaborative.org/) - a type of interactive decision tree - that guides the assessor through the assessments and helps with identifying the most suitable section and provides the classifications after answering a set of questions. The calculator additionally provides a printout of the assessment process with displaying information collected during the assessment and the overall classification of the variant.

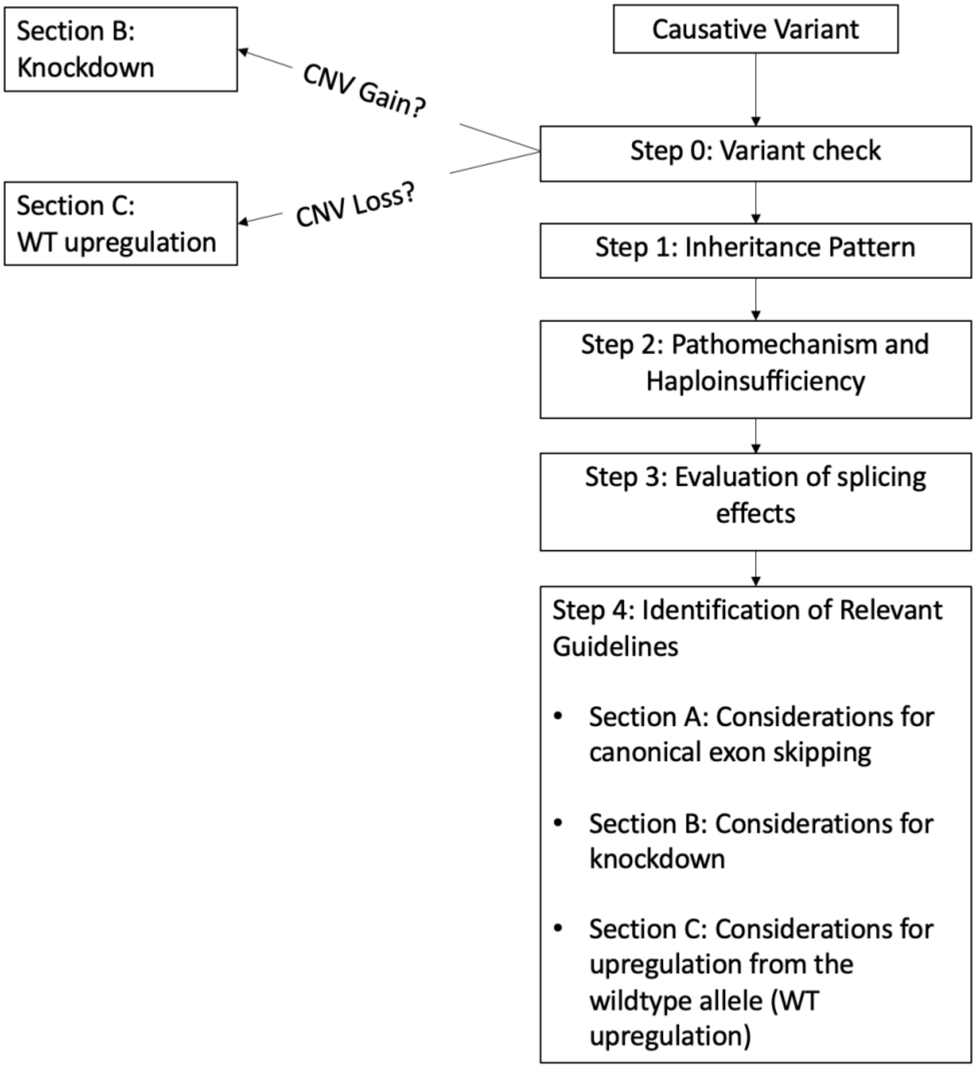

**Figure 4: Overview of consensus guidelines document.**

The guidelines begin with Step 0, 1, and 2, which highlight the assessment of the variant description, inheritance pattern, and pathomechanisms of the variant. When applicable, the evaluation of splicing effects is considered in Step 3. Dependent on the information gathered in Steps 0-3, readers are directed to Sections A, B, or C. The document is designed in a way in which assessors are directed to relevant sections, and are not required to read the entire document for assessment of a variant.

### Step 0 - Variant check

In this step, the assessor will learn which type of genetic variant can be assessed using these guidelines, and how to ensure that the variant description provided is correct. Without the correct variant description, an assessment is not possible.

Variants applicable to these guidelines are restricted to (likely) pathogenic, disease-causing single nucleotide variants, small indels, one or multi-exon deletions or duplications, and single gene deletions or duplications. Currently excluded are variants in non-coding genes, deletions/duplications spanning multiple genes (i.e., contiguous gene syndromes), imprinting defects/uniparental disomy, structural rearrangements (e.g., translocations), and aneuploidies. Additionally, mitochondrial DNA disorders (i.e., variants in the mitochondrial genome) cannot be evaluated with these guidelines. All variants that cannot be evaluated will be classified as "unable to assess". Insertions within coding regions are mostly not applicable unless confined within an exon that can be skipped; insertions in introns can be applicable, one example being milasen (Kim et al., 2019). Repeat expansion disorders can also be assessed using these guidelines but the pathomechanism should be well understood to evaluate for knockdown approaches or exon skipping approaches.

Before starting to assess a variant, it is important to check the accuracy of the variant description. That means, is the gene symbol correct, does the transcript match the variant and is the consequence on protein level denoted accurately? If the variant description is incorrect, the variant should be classified as "unable to assess" and the description be corrected and verified. These guidelines work with the MANE select transcript but apply to any other transcript also. Choosing a different transcript when it has more biological relevance in a given disease context is preferred.[1]

To check a variant and its description, please follow the HGVS nomenclature. Correct variant description and matching of gene and transcript can be checked with Mutalyzer and VariantValidator. Note, Mutalyzer cannot normalize intronic variants given for a non-genomic reference sequence, i.e., an NM accession number. Examples of variants and their descriptions are provided at the end of these guidelines.

**Considerations for Single Gene Copy Number Variants (CNVs)**

For the gain or loss of whole genes, special considerations apply. For the gain of a whole gene, knockdown strategies should be considered (see Section B). For the loss of a gene with one functional wildtype copy left, upregulation from the allele should be considered (see Section C). ASO strategies cannot be applied when no copies of the gene are present, i.e., loss of both gene copies or loss of one gene copy for hemizygous genes.

[1] In some instances, variants that are deep intronic in the MANE select transcript are exonic in another transcript and are disease-causing due to the effect they have in that exon. Please pay attention to this exception when evaluating the functional consequences of a variant.

***Optional ASO Check After a Variant Check***

To save time for assessors familiar with these guidelines, we recommend checking whether there has already been an ASO/siRNA developed for the variant in question (clinically or pre-clinically) immediately after checking the correct variant description. This can mean that an ASO has been developed for the specific variant, e.g., a splice correction ASO or a gapmer ASO, or an ASO has been developed for an exon skipping approach for an exon this variant is located in. It can also be that a gapmer ASO/siRNA is available for the gene in question or allele-specific for a SNP that is in phase with the pathogenic variant. When doing this ASO-availability check, it should be ensured that the ASO strategy identified also applies to the variant under assessment. If in doubt, we strongly suggest following the full guidelines and at the end checking for available ASOs. Generally, if an ASO has been developed, it should be carefully evaluated whether there is enough functional evidence that the ASO development was successful. That means, for example, demonstrating restoration of protein levels or rescue of a cellular phenotype. In cases where there is enough functional evidence, the variant can be classified as "eligible", and no further evaluation is necessary[2,3]. To investigate whether an ASO has already been developed, we recommend a thorough search using ClinVar, Pubmed, Google Scholar, and web search, also paying attention to conference abstracts if available. Further, ASO registries are available (e.g. n-Lorem) that can be accessed to identify available ASOs. We also consider it sufficient if there is an ASO/siRNA already in clinical implementation, even if there is not yet published data available. That means if an ASO is available in the registries and has already been administered to one or more individuals, this is sufficient to classify the variant under assessment as "eligible".

---

[2] Please note that if publications are available that test ASOs for a specific variant/exon/gene, one has to carefully read whether the ASO design and development was indeed successful. It could be that ASOs were designed but the transcript or protein level could not be restored. Now it depends whether it might not be possible at all to, for example, skip a canonical exon, or whether another ASO design approach might still be justifiable. Depending on the assessment, that might lead to a "not" for eligibility or a "likely/unlikely" following the guidelines. We generally recommend that at least two independent groups with sufficient functional evidence should have shown that ASO development is not possible to declare a variant "not eligible".

[3] In the case of an allele-specific ASO development, please check whether the ASO was developed for the specific variant or for a SNP. For the latter, only if the patient has that exact SNP in phase with the pathogenic variant would that ASO be applicable for that patient. If this is not the case, proceed with the next steps of the assessment.

### Step 1 - Assessment of pattern of inheritance and disease type

In this step, assessors will identify the pattern of inheritance of the variant under assessment and the disorders implicated in the disease gene. Understanding the inheritance pattern of the variant and the diseases associated with a gene is crucial to later decide on the most suitable ASO strategy and thus the section to use for the assessment. Different considerations will apply depending on the inheritance pattern.

The assessor has to identify if the variant is inherited in an autosomal dominant (AD), autosomal recessive (AR), or X-linked manner. Usually, in the case of an (autosomal) recessive inheritance, the variant is either being reported as homozygous or a second variant in trans has been identified (i.e., compound heterozygous), making this step of the assessment straightforward. In the case of X-linked disorders in XY males, the variant will be hemizygous. For (autosomal) dominant inheritance, one variant should be reported either *de novo* or inherited from one of the parents. Should the respective information not be available (e.g., the phase of two variants in an autosomal recessive disease gene), some other steps can be taken to gather the necessary knowledge. The following websites can be used to identify the inheritance pattern of a disease:

OMIM https://www.omim.org/
Orphanet https://www.orpha.net/consor/cgi-bin/index.php
GeneReviews https://www.ncbi.nlm.nih.gov/books/NBK1116/
Pubmed https://pubmed.ncbi.nlm.nih.gov/
ClinGen https://www.clinicalgenome.org/

Some genes are implicated in different diseases which can, for example, have an autosomal dominant and recessive pattern of inheritance respectively. In such instances, it is important to identify which pattern of inheritance applies to the specific case. A web search on the variant can be useful. The variant may have been reported in a publication where a pattern of inheritance is noted. Also, gnomAD can be of help. The population frequency of a variant can indicate if the variant is associated with a dominant or recessive inheritance. For example, if a LoF variant has a high allele frequency for heterozygotes but no homozygotes are reported, the variant is more likely to be associated with an AR pattern of inheritance. Similarly, for X-linked disorders in XY males, a lack of hemizygotes would be the equivalent assessment. Further, checking with the diagnostic laboratory or the treating clinician and reviewing the family history can be useful to gather more information on the inheritance pattern in that specific case.

Should the gene be implicated in different diseases with distinct inheritance patterns, we recommend noting this down as this can help with later steps of the assessment. For example, a gene associated with an AD disorder caused by heterozygous GoF variants and also associated with an AR disorder caused by LoF variants implies that targeting the GoF variants with a knockdown approach is possible but loss of too much of the gene product will also be detrimental.

### Step 2 - Assessment of pathomechanism of the genetic variant and haploinsufficiency

In this step, assessors will identify the pathomechanism of the variant and assess whether the gene in question is associated with haploinsufficiency. The pathomechanism is relevant to decide on the most applicable ASO strategy and with this, the sub-guidelines to use for the assessment. Identifying whether a gene is associated with haploinsufficiency is crucial to deciding whether ASO approaches need to be allele-specific and whether the assessors should consider the guidelines on upregulation from the wildtype allele (see Section C).

A pathogenic variant can lead to different effects. The variant can lead to a loss of function of a protein (LoF), toxic gain of function (GoF), or dominant-negative (DN) effect. For an explanation of the different pathomechanisms, please see, e.g., Backwell & Marsh, 2022.

Assessing the variant effect can mostly be done by conducting a web search and reading up on publications and reports of the variant. Resources that can help in identifying the pathomechanism associated with a variant are:
OMIM https://www.omim.org/
Orphanet https://www.orpha.net/consor/cgi-bin/index.php
GeneReviews https://www.ncbi.nlm.nih.gov/books/NBK1116/
Pubmed https://pubmed.ncbi.nlm.nih.gov/

Note that for many variants, no functional studies have yet been performed, especially if the variants are very rare. Thus, we list here some considerations on how to define the pathomechanism:

- Variants with a predicted LoF effect like nonsense and frameshift variants can be assumed in many clinical contexts to result in a null allele in the absence of functional studies (Abou Tayoun et al., 2018). Note that exceptions apply. For example, nonsense or frameshift variants leading to a premature stop codon in the last exon or within 50 bp of 3' end of the penultimate exons may not necessarily lead to loss-of-function. In this case, assessors should take into consideration protein domains and presence of downstream (likely) pathogenic variants (Abou Tayour et al., 2018). In a disease that has only been associated with LoF variants, a newly reported likely pathogenic or pathogenic missense variant may be LoF, especially if this variant is in trans with a known pathogenic LoF variant (for recessive diseases).[4]
- For genes where both LoF and GoF are a known cause of disease, assessing the pathomechanism of a missense variant is challenging. Here, it will become important to take the phenotype and inheritance pattern of the variant into account to make a decision on the pathomechanism of the variant. It can, for example, be possible to predict a pathomechanism in cases where LoF and GoF variants lead to distinguishable

[4] We recommend restricting these assumptions to AR diseases because in some cases nonsense and frameshift variants can cause splice aberrations and ultimately lead to a GoF effect on protein level (Flanagan et al., 2017).

phenotypes. There are only rare cases where phenotypes are distinct enough to make such a decision. One such example includes distinctly different phenotypes associated with GoF and LoF variants in *GABRB2* patients (Mohammadi et al., 2024). On the other hand, in cases where LoF and GoF variants lead to similar phenotypes as seen in intellectual disabilities, more evidence is necessary and functional studies are crucial. Generally, when in doubt, functional evidence should be obtained.

- DN variants are, by definition, dominant and can thus only be found in dominant disorders. However, distinguishing whether a variant is GoF or DN might be difficult. Fortunately, for GoF and DN variants, the ASO approaches are mostly the same.
- There are rare reports of homozygous GoF (Schwarz et al., 2020) variants, thus carefully checking the inheritance pattern and pathomechanism is important.
- A missense variant might cause the loss/disturbance of an inhibitory domain causing a toxic gain of function effect of the protein (Mohassel et al., 2021).

If sufficient functional evidence does not exist for a given variant, the next step would be to request more information on the variant or experimentally determine the pathomechanism. In the meantime, classify the variant as “unable to assess”.

#### Haploinsufficiency

Besides the identification of the inheritance pattern and the pathomechanism of a variant, the associated gene also needs to be evaluated for haploinsufficiency. Haploinsufficiency refers to a situation in which one healthy, wildtype allele does not generate sufficient protein product to preserve the physiological state (Deutschbauer et al., 2005). In most cases, haploinsufficiency is connected to AD disorders associated with LoF variants; however, several genes have been identified that are associated with LoF variants causing haploinsufficiency, and also GoF variants. That is the case for example for the genes *SCN2A* and *SCN8A* (Li et al., 2021; Wagnon et al., 2017). Here, GoF and LoF variants cause different clinical presentations depending on the pathomechanism of the variant.

The knowledge of whether a gene is associated with haploinsufficiency is important to decide on the best therapeutic approach to assess a variant (see Fig. 6). In brief, for AD disorders caused by LoF variants, in case of haploinsufficiency, the healthy wildtype allele could be upregulated using an ASO strategy (see Section C). For GoF variants, it will be important to assess whether an allele-selective approach is necessary for transcript knockdown (see Section B).

To determine whether a gene is associated with haploinsufficiency, different resources can be used. Often, a web search can help as well as GeneReviews.
GeneReviews https://www.ncbi.nlm.nih.gov/books/NBK1116/
Pubmed https://pubmed.ncbi.nlm.nih.gov/

Further indications of whether a gene is associated with haploinsufficiency can be gained from checking the gene constraint metrics like the pLI and LOEUF scores in gnomAD, or the haploinsufficiency score determined by the ClinGen consortium. Additional information on dosage sensitivity in general and how one can assess dosage sensitivity is provided in Section B. We consider a curation from a reputable independent source, such as the ClinGen consortium, the highest level of evidence for dosage sensitivity/haploinsufficiency.

### Step 3 - Evaluation of splicing effects

In this step, assessors will evaluate the effect on splicing of each variant. As explained in the Background, correction of aberrant splicing is an elegant way to restore the reading frame and the physiological splicing pattern (Fig. 1). Thus, the first aim should be to assess whether a splice-switching ASO strategy is applicable. Whenever this is not a possibility, the flowchart in Step 4 will aid in choosing the most suitable section for variant assessment. For the splicing evaluation and correction of aberrant splicing, different considerations have to be made given inheritance patterns and pathomechanism of the genetic variant (described separately at the end of Step 3). Table 3 provides an overview of the classification of variants for eligibility to ASO splice correction.

In this section, the following is covered:

1. Determining whether a variant affects splicing and what is considered sufficient evidence for mis-splicing
2. Considerations for eligibility of splice correction ASOs for both intronic and exonic variants
3. Types of exonic variants which can cause aberrant splicing and alternate ASO strategies to consider in place of splice correction
4. Important considerations for pathomechanism and inheritance pattern
5. How to search the literature for splice correction ASOs

Different types of variants can influence the splicing process and thus the decision on which ASO strategy to apply. While all variants should be assessed for their splice-altering potential (Anna & Monika, 2018), some exceptions usually do not have to be evaluated in Step 3:

- For nonsense and frameshift variants that are associated with a LoF mechanism at the protein level, see Section A[5] (or Section C in cases of haploinsufficiency). Rare cases where nonsense and frameshift variants that affect splicing lead to GoF or DN effects can be assessed with the considerations outlined in Step 3.
- For whole exon duplications and deletions, please consider the variant effect and jump to Section B (knockdown) or Section C (upregulation from the wildtype), check Fig. 6 for directions.

The effects on splicing of each variant need to be confirmed with a functional assay, whereby only RNAseq, qPCR, or cDNA sequencing/analysis obtained from patient-derived cells can be considered sufficiently reliable. Please note that it is considered sufficient if functional data of the above-mentioned kind is available on the same variant from a different patient, i.e., a case report in the literature or information provided in ClinVar. Data gathered through mini- and midi-genes cannot be considered sufficient as these assays do not take the full genetic environmental context into account and results can be misleading (Lin et al., 2021). Explicitly, the prediction of splicing effects using *in silico* tools is not sufficient evidence and cannot be used for these assessments (Oh et al., 2024).

[5] One notable exception applies here. Nonsense and frameshift variants have a small theoretical probability of resulting in aberrant splicing (Haque et al., 2024). Depending on the splicing effect, this could be canonical exon skipping, in which case the variant is not eligible for exon skipping treatment. It could also be partial exon skipping/cryptic splicing in which the exon in which the variant is located can still be considered for canonical exon skipping, see Section A.

In case there is functional evidence against a splice-altering effect, i.e., confirmation that the variant does not affect splicing, please refer to Fig. 6 to guide you towards the next section applicable to your case. Additionally, if there is no functional evidence of splicing effects, please refer to Fig. 6. In the case where there is no evidence of splicing effects (whether for or against), this does not mean the variant is ineligible for splice correcting ASO. A lack of functional evidence towards splicing means this variant cannot be assessed for slice correction eligibility at this time. Theoretically, all variants are suspicious of splicing until demonstrated otherwise, and if evidence on splicing effects becomes available, the variant should be reassessed for eligibility towards splice correction ASOs.

If splicing effects are confirmed, the exact effects on splicing need to be evaluated. This can be a gain of an acceptor or donor splice site or the loss/weakening of an acceptor or donor splice site. Especially, if canonical splicing is destroyed - due to canonical splice site variants, branchpoint variants, or variants destroying other splice-regulatory elements[6] - the variant is most likely not eligible for a splice correction ASO treatment. Canonical splicing is considered destroyed if no wildtype transcript/splicing at the canonical splice sites can be identified in the functional analysis. If some wildtype transcript is still produced or protein function detected, canonical splice sites are considered as weakened. Variants that fully abolish canonical splicing are not amenable to a splice correction ASO and are classified as “not eligible” for this approach.[7,8]

We recommend studying available data carefully and to also assess data provided in supplementary material, as one can often find relevant gels and blots in the supplement that may only be hinted at in the main manuscript. Further, look out for evidence illustrated by gels or qPCR results etc. instead of solely relying on how the authors describe or discuss the data. From our experience, some manuscripts claim there is no wildtype splicing left whereas faint wildtype bands can be identified in a gel, which might be sufficient to consider this a suitable case. Please consider that a variant's effect on splicing may not have been assessed. The absence of evidence might indicate that splicing effects were overlooked.

We generally distinguish intronic and exonic variants for the splicing assessments. Please also see Fig. 5 for a general overview of classification of variants towards splice correction.

**Intronic variants**

As a rule of thumb, we consider intronic variants that are >-100 bp (upstream of acceptor splice site) or >+50 bp (downstream of donor splice site) away from the nearest canonical splice site as likely eligible, i.e., there should be no negative impact on the canonical splice sites and branchpoint (Fig. 5). For variants closer to the canonical splice sites, enough functional evidence

[6] Under certain circumstances, aberrant splicing caused by a destroyed splice enhancer can be counteracted by the steric blocking of a splice silencer. The regulatory element to be blocked should be strong enough and react in *cis* to the affected splice-regulatory element.

[7] Two notable exceptions apply here. If the destruction of canonical splicing leads to exon skipping of an out-of-frame exon, skipping of adjacent exons to restore the reading frame can be considered but criteria outlined in Section A apply. If the destruction of canonical splicing leads to a partial skipping of the exon (a few bases are missing - usage of cryptic splice sites) resulting in a frame-shift, ASO-induced canonical exon skipping can be considered for exons that fulfill criteria outlined in Section A.

[8] The assessment of whether the wildtype allele is still left can be difficult. For homozygous and hemizygous variants, this is straightforward, but in the case of heterozygous or compound heterozygous variants, this analysis is more difficult and information on the other allele is necessary to assess the effect of a variant.

should be available on the effect on canonical splicing. Variants within 5 bp of the exon-intron/intron-exon boundary are considered “not eligible” for splice correction.[9] Note that the ASO should also not disturb the branchpoint, which can vary and is usually found 18-40 bp upstream from the acceptor splice site, but can be in 10-100 bp upstream (i.e., -10 to -100 bp region) (Xie, Wang, & Lin, 2023).

Intronic variants with functional evidence of causing aberrant splicing can now be analyzed using the above-mentioned criteria and classified using Table 3.

**Exonic variants**

For exonic variants, further considerations apply. As for the intronic variants, the exonic variant should be at least 5 bp outside of the canonical splice sites (upstream and downstream - hard cut-off). We further define a second cut off at 15 bp (region 6 to 15 bp upstream or downstream of the canonical splice sites) as a soft cut off where a splice correction can be considered but is challenging. This cut off is based on the idea that ASOs can bind to this region without destroying the canonical splice set, yet there is still a possibility of weakening canonical splicing.

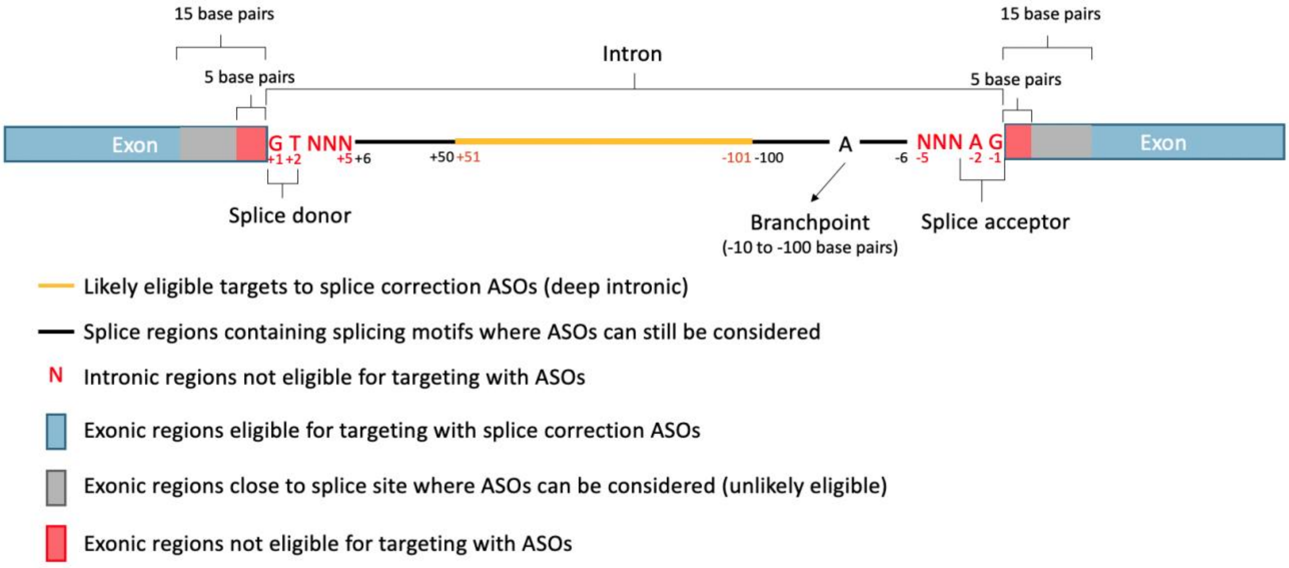

**Figure 5: Eligible targets for splice correcting ASOs**

Design of splice correcting ASOs takes into consideration key splicing motifs. The most amenable variants for splice correction are deep intronic variant, highlighted yellow in this figure. As the canonical splice sites are approached, one must consider the effects of the ASO blocking important splice site motifs (i.e., the branchpoint). These regions are highlighted black (intronic) or gray (exonic) in this figure. Anything within 5 bp is considered not eligible for targeting with an ASO (highlighted red in this figure, both intronic and exonic).

[9] Avoiding +/-5 bp around the exon-intron/intron-exon boundaries is recommended because these variants are most likely destroying canonical splicing (or the splice site itself) which cannot be corrected. Additionally, it will be challenging to place an ASO within this region without negatively impacting canonical splicing. However, there are currently ongoing research efforts that aim at identifying ways to counteract aberrant splicing caused by variants within this region. Should this approach become feasible in the future, the guidelines will be adjusted.

For exonic variants, the specific type of variant is important for further analysis.

1. Nonsense and frameshift variants that cause aberrant splicing can be further distinguished:
    a) If the splice aberration itself leads to a LoF effect on the protein, the variant cannot be analyzed using this part of the guidelines. Correction of splicing would still lead to a LoF on protein level. Thus, these variants fall under Section A for exon skipping analysis (or Section C in the case of haploinsufficiency).
    b) If the splice aberration leads to a toxic GoF effect on protein level, the variant can be considered for splice correction. Correction of the splicing effect will then lead to an early truncation and most likely a LoF effect on protein level. For certain cases, this is a useful ASO approach. It is also possible to consider these variants for downregulation (Section B).

    In both situations our considerations for haploinsufficiency should be taken into account.
2. Synonymous variants that cause aberrant splicing do not influence the amino acid sequence and can be assessed using Table 3.
3. For missense variants and small in-frame indels (<50 bp) (Mahmoud et al., 2019), the considerations are more complicated as both the effect on splicing and also the effect of altering the protein coding sequence need to be taken into account. It needs to be established that the variant effect at the protein level solely arises from the aberrant splicing and not that a missense variant or small indel itself causes a pathogenic effect. It is possible that a missense variant causing aberrant splicing leading to a LoF at the protein level, could independently cause a GoF or DN effect as a missense variant.
    a) If the effect of a missense variant or in-frame indel on the protein sequence is not known (independent of the splice-altering effect), the variant cannot be assessed until further evidence is available[10]
    b) If the change in amino acid sequence is known to be pathogenic, for example, if a different pathogenic nucleotide change causes the same amino acid change without the splicing effect, different sub-criteria apply:
        i) Aberrant splicing and missense variant cause GoF/DN effect → can be considered for downregulation (Section B) or for exon skipping (Section A)
        ii) Aberrant splicing and missense variant cause LoF effect, with the amino acid change causing a complete loss of function on protein level → can be considered for exon skipping (see Section A)
        iii) Aberrant splicing causes GoF effect and missense variant would lead to a LoF → can be considered for exon skipping (Section A) or knockdown (Section B) and also for splice correction if the LoF phenotype is milder or loss of one allele is tolerated
        iv) Aberrant splicing causes LoF effect and missense variant causes GoF/DN effect → can be considered for exon skipping (Section A) or for splice

[10] One can consider exon skipping for the variant independent if the variant is LoF, GoF, or DN based on the amino acid change if it is possible to show that the resulting protein has some remaining typical function. Please see Section A.

correction in case of a GoF effect that leads to a less severe phenotype compared to the LoF phenotype

v) Aberrant splicing leads to GoF or LoF effect and the variant causes partial loss of function/reduced protein function → can be considered for splice correction and would be classified as "unlikely eligible" for splice correction. Here, the underlying thought is that having a partially functional protein is better than having no protein function or toxic protein function due to aberrant splicing. Variant can also be considered for exon skipping (Section A) or for downregulation (Section B) in case of GoF effects from aberrant splicing.

c) If the amino acid change is known to be benign on protein level, due to population data or functional studies, the variant follows the same considerations as a synonymous variant and can be analyzed using Table 3.

**Important considerations for different inheritance patterns and pathomechanisms**

For AR disorders, Step 3 applies without restrictions. If the variants are homozygous or compound heterozygous, the Step 3 evaluation has to be done once or twice for both variants. If the disorder is AD associated with a LoF, the Step 3 guidelines also apply. Since the purpose of these ASOs is to restore wildtype splicing, rather than alter splicing, whether the ASO binds the wildtype or pathogenic allele does not matter (unlike with canonical exon skipping ASOs and gapmer ASOs). Similarly, variants that lead to a GoF or DN effect through altering splicing can be assessed with these guidelines (please consider the effect of variant on protein level once splicing is corrected). For GoF or DN variants, however, one might consider a knockdown approach for a more efficacious effect (see Section B).

**ASO check**

In addition to the recommended assessment strategies, assessors should review the literature for splice correcting ASO strategies. This review can be performed either as the final step to validate the assessment strategy or earlier in the assessment process (see Step 0). Specifically, for splice correcting ASOs, it is crucial to evaluate whether a splice correcting approach has been implemented and validated for the specific variant at the RNA level. Please also consider that the rescue at RNA level needs to be sufficient to produce enough protein to rescue the phenotype. Ideally, a publication should provide this evidence.

Splicing mechanisms differ by variant, making it crucial to ensure that any existing ASOs found in the literature are applicable to the specific variant of interest. Additionally, if the literature indicates that an ASO is ineffective in correcting splicing for the variant of interest, this does not necessarily render the variant ineligible for ASO development. We recommend that a variant should only be considered "not eligible" for splice-correcting ASOs if there is evidence from two independent investigations at the protein or functional level, or from one investigation providing a convincing explanation as to why an ASO cannot be developed.

In cases of conflicting evidence, consider the quality of the research, the types of experiments conducted, the nature of the results shared, and the publication date. The evaluation of literature on existing ASOs should be carried out at the assessor's discretion, with a critical and discerning approach.

To help with identifying available ASOs for splice correction, we recommend a search term in Pubmed like this (searching for an ASO targeting a specific variant), text in bold would need to be adjusted for the gene and variant being examined:
***ABCA4*** *AND ((ASO) OR (AON) OR (antisense oligonucleotide)) AND ((***Gln876Ter***) OR (***c.2626C>T***) OR (***E876X***) OR (***Q876X***) OR (***Gln876****) OR (***E876****) OR (***Q876****))*

**Table 3: Classification of variants for their eligibility towards splice correction.**

| Classification | Criteria |
|---|---|
| Eligible | ASO has already been developed and shown to work with available functional evidence at the protein level (pre-clinical data is sufficient) |
| Likely eligible | **Functional studies (RNAseq, qPCR)** validate alternate splicing<br><br>**AND (if intronic)**<br><br>{<br>Intronic variant -101 bp or +51 bp outside of the canonical splice sites<br><br>**OR**<br><br>No weakened branch point/canonical splice site as determined by functional studies<br>}<br><br>**AND (if exonic)**<br><br>Donor/acceptor gained is not within 15 bp of a canonical splice site<br><br>**AND (if exonic)**<br><br>Functional evidence shows there is no pathogenic effect of the amino acid change(s) on the protein. |

| | |
|---|---|
| Unlikely eligible | **Functional studies (RNAseq, qPCR)** validate alternate splicing<br><br>**BUT**<br><br>{<br>Canonical splice site and branchpoint is weakened (but still functional, i.e., either canonical transcript or protein function still detectable)<br><br>**OR/AND (if intronic)**<br><br>Intronic variant within -6 and -100 bp or +6 and +50 bp of the canonical splice sites.<br><br>**OR/AND (if exonic)**<br><br>Donor/acceptor gained within 6-15bp of the canonical splice site.<br><br>**OR/AND (if exonic)**<br><br>There is evidence of residual protein function (i.e., if splicing is corrected, the protein coding change still produces a partially functional protein).<br>} |
| Not eligible | Canonical splice site and branchpoints are destroyed<br><br>**OR**<br><br>Different nucleotide change leading to the same predicted amino acid residue change – but no alternate splicing – is pathogenic<br><br>**OR**<br><br>Variant within 5 bp of the canonical splice site<br><br>**OR**<br><br>Evidence that ASO cannot be developed, shown by two independent investigations on the protein/functional level **or** one investigation with a convincing explanation of why ASO cannot be developed. |

### Step 4 - Identification of Relevant Guideline

In this step, assessors will be guided towards the sections (sub-guideline) applicable to the variant under assessment. Once the inheritance pattern, pathomechanism, and splicing effects are evaluated, a decision on the most useful guideline(s) can be made. If a variant does not have an effect on splicing, or an effect on splicing that cannot be assessed using Step 3, the assessor can turn to the following flow diagram and chart for assistance.

The table within the flow chart lists all applicable sections and with the knowledge of the inheritance pattern and pathomechanism, the assessor can now identify the applicable section(s) to be read to make the assessment.

Please note again that each section is a stand-alone sub-guideline and thus one can jump to the necessary section without having to read the full text.

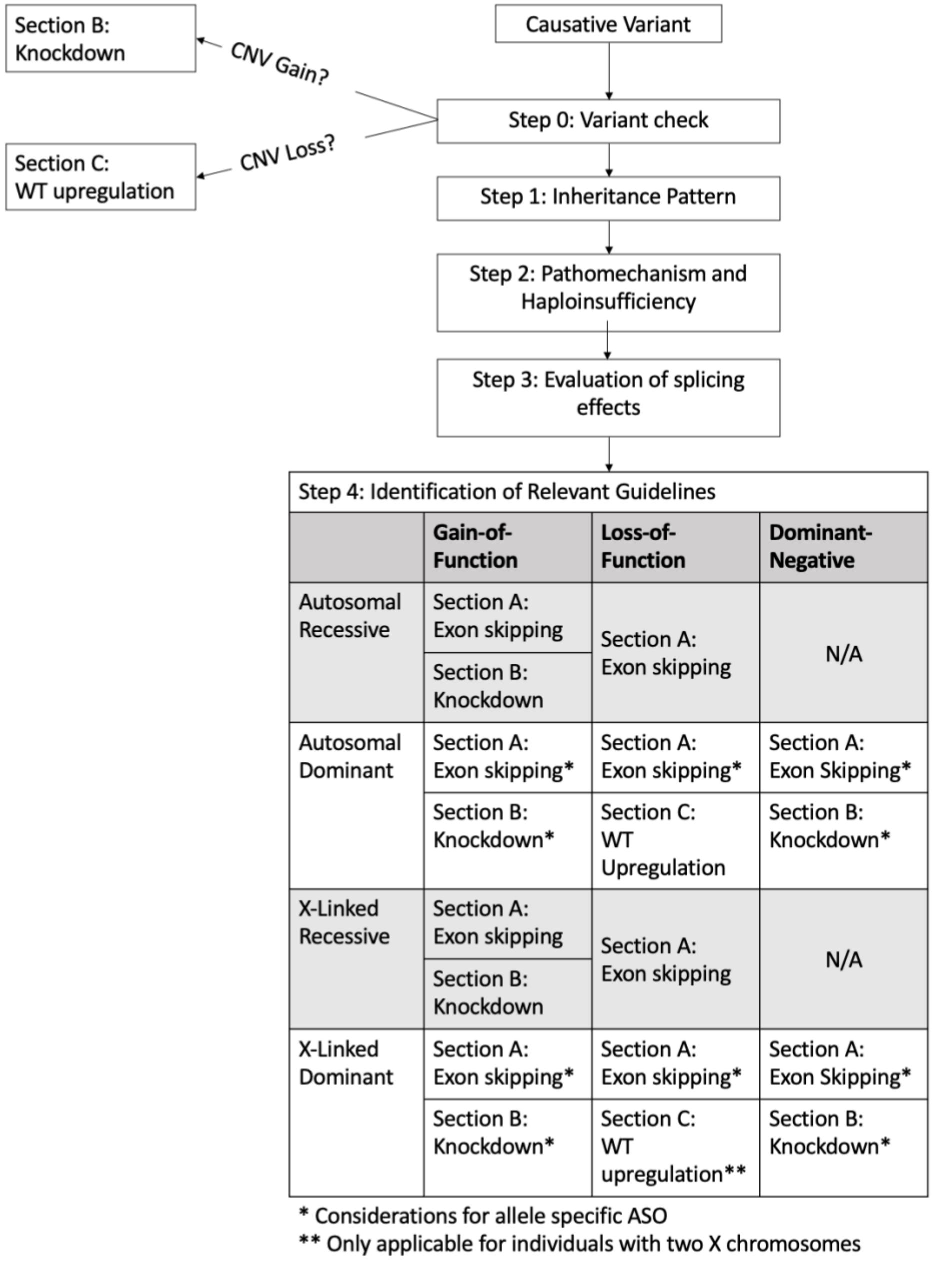

**Figure 6: Flowchart for the identification of relevant guidelines.**

Steps 0, 1, and 2 require the assessment of the variant description, inheritance pattern, and pathomechanisms. The evaluation of splicing effects is considered in Step 3. Depending on the information gathered in Steps 0-3, one can consider the guidelines discussed in Sections A, B or C.

### Section A - Considerations for Canonical Exon Skipping

In this section, assessors will be guided on how to assess a variant for eligibility towards canonical exon skipping ASOs. As described in the Background, exon skipping ASOs can be used to skip exons containing the pathogenic variant (Fig. 1). Variants generally to be considered for exon skipping are nonsense and frameshift variants causing a LoF in AR, AD, and X-linked disorders (additional considerations apply in dominantly inherited diseases, see “Important considerations for different inheritance patterns and pathomechanisms” at the end of this section). GoF and DN negative variants can also be amenable for an exon skipping approach under certain circumstances as well as whole exon deletions (see “Important considerations for different inheritance patterns and pathomechanisms” in this section). While splicing assessments are based on the variant, evaluating the potential for exon skipping are based on the exon, thus <u>an exon skipping approach is applicable to all variants within that exon</u>. For an exon to be skippable, different criteria need to be met (Fig. 7).

This section covers the following topics:

1. Assessing exon position and frame
2. Assessing exon size
3. Strategies for searching and considering naturally occurring exon skipping and in-frame deletions
4. Assessing the role of functional domains
5. Important considerations for different inheritance patterns and pathomechanisms
6. Strategies for searching the literature for exon skipping ASOs

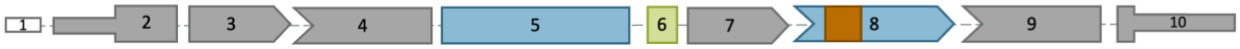

**Figure 7: Overview exon skipping assessment using a hypothetical transcript.**

The first and last coding exons (exon 2 and 10) cannot be skipped, in addition to out-of-frame exons (exons 3,4,7,9, shown by the shape of the exons). In-frame exons encoding for more than 10% of the coding region (exon 5) and exons coding for functional domains (exon 8) are unlikely eligible for exon skipping. Likely eligible for exon skipping are small, in-frame exons that do not code for a domain (e.g., exon 6). White: exon not considered for canonical exon skipping, e.g., exons in the UTRs, grey: Exons not eligible for skipping, blue: exons unlikely eligible for exon skipping, green: exon likely eligible for exon skipping, orange: functional domain.

***Assessing Exon Position and Frame***

The first and last coding exon are usually not eligible for exon skipping as this would lead to the loss of the start or stop codon. Note, due to untranslated regions (UTRs) the first and last exons of a transcript are not necessarily coding exons and, thus, assessors should be aware of which exons contain the canonical start and stop codons (Aspden, Wallace, & Whiffin, 2023) (Fig. 7). Additionally, genes containing exactly one coding exon are not eligible and are disqualified from further analysis. Next, assessors should evaluate the length and position/frame of the exon. Out-of-frame exons (containing a number of base pairs not divisible by 3) should be classified as “not eligible” in the case of LoF variants where a restoration of the reading frame is the aim. Assessors can use tools such as the UCSC Genome Browser, ExonViz (van den Berg, Lauffer, & Laros, 2024),

or Ensembl to determine whether the exon is in-frame or out-of-frame and whether the exon is coding or non-coding (Fig. 8).

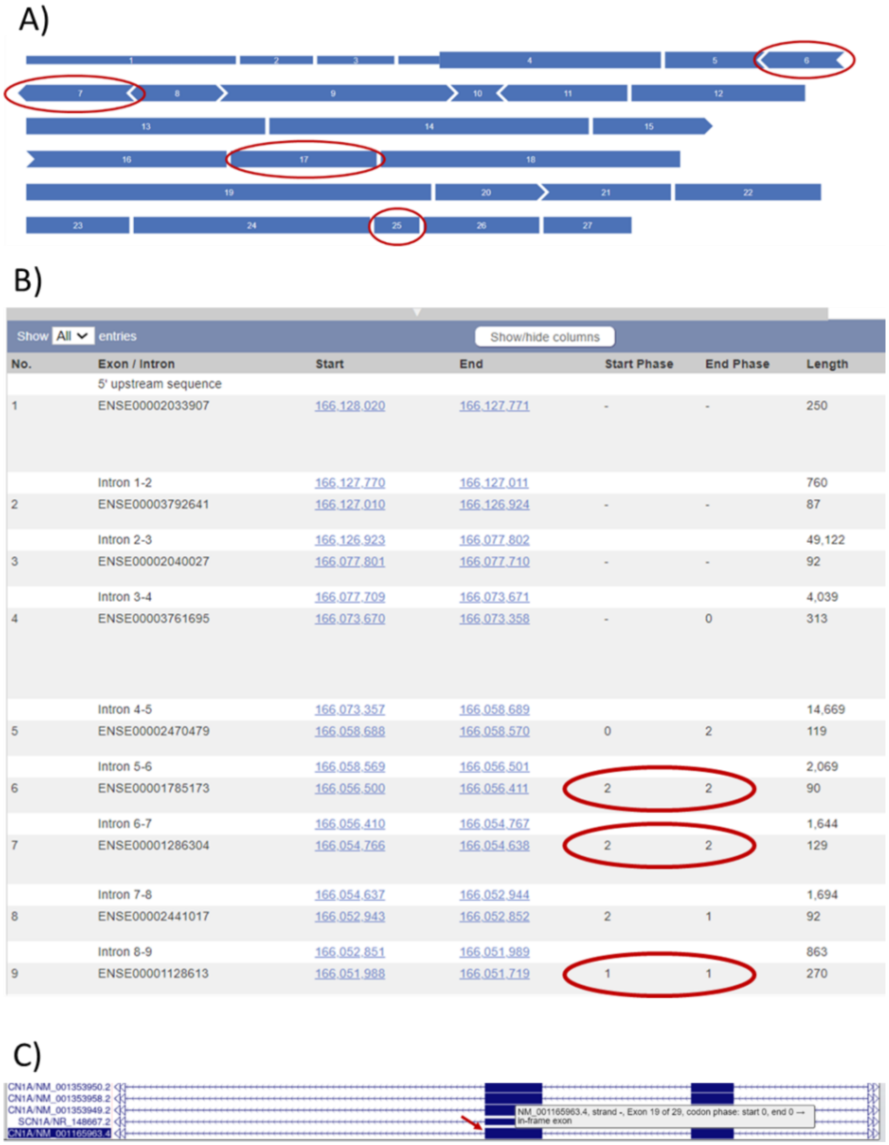

**Figure 8: Determining exon frames.**

**A)** Exon frames in ExonViz can be identified by their shape. Rectangular shapes are in-frame with a phase at 0-0 (exon starts and ends with the codon). Exons with an arrow on one end and a notch on the other end are also considered in-frame, whereby it does not matter if the arrow is upstream or downstream. If there are two arrows or two notches, the exons are out-of-frame. An arrow indicates a 1 nucleotide overhang while a notch illustrates a 2 nucleotide overhang. Small, in-frame exons are circled in red (exon 6, 7, 17 and 25). For example, exon 6 starts with an arrow, meaning the exon starts with 1 nucleotide from a codon starting in the previous exon (i.e., this is the third nucleotide in the codon, with the first 2 nucleotides being found in exon 5). Exon 6 ends with a notch, indicating that these are the first two nucleotides of a codon (2-2 phase). In total, both ends together cover 3 nucleotides. **B)** Identification with Ensembl can be done by checking the phases of an exon. Exons with phases 0-0, 1-1, 2-2 are in-frame. Additionally, dividing the exon length by 3 can help determine the frame. **C)** UCSC Genome Browser shows phases to determine the frames using mouse over. The browser directly indicates the phase for you. Please ensure you are hovering over the correct transcript.

For in-frame exons, it is possible that the first and last codon is partially encoded by the adjacent exons (exons with phases 1-1 and 2-2). In this case, the formation of a stop codon is possible at the new exon-exon boundary created by splicing the upstream and downstream exon together. Assessors should evaluate whether joining the adjacent exons would form an in-frame stop codon, coded by either a TAA, TAG, or TGA (Fig. 9). If this is the case, the exon is not eligible for exon skipping (exceptions apply, please see below). Assessors can utilize the UCSC Genome Browser or Ensembl to check (instructions on how to perform this check are provided in video 11, the NM_000170.3(GLDC):c.538C>T example).

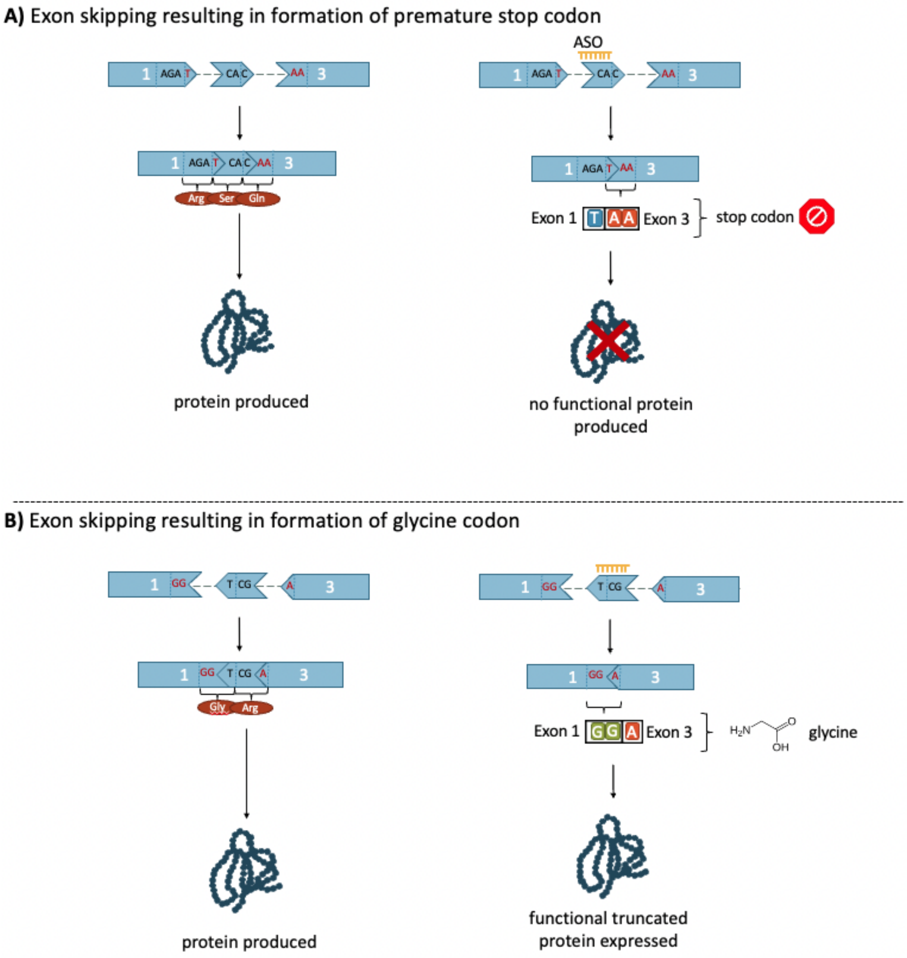

**Figure 9: Formation of a new codon as a result of exon skipping.**

When an in-frame exon with phase 1-1 or 2-2 is skipped, the adjacent exons will form a new codon on the new exon-exon boundary. Panel A shows the formation of a stop codon when an ASO is used to skip in-frame exon 2. The “T” nucleotide from exon 1, and “AA” nucleotides from exon 3 join together to form a stop codon. This leads to a premature termination resulting in an absent gene product or a truncated, non-functional product. Panel B shows the formation of a new codon with a functional product. The “GG” nucleotides at the end of exon 1 join the “A” nucleotide at the start of exon 3, upon skipping of exon 2. This codes for glycine.

The skipping of an exon that disrupts the reading frame (skipping out-of-frame exons, forming premature termination codons, or skipping the first and last coding exon) tends to be not eligible for analysis. However, there are exceptions to these rules:

- It is theoretically possible to skip out-of-frame exons to generate a premature termination codon within the last or penultimate exon given that there is some left-over function of the protein (please note that this is theoretically possible - one would still have to assess the last exon for the importance of domains and whether this shortened transcript is predicted to undergo nonsense mediated decay)
- It is also possible to skip out-of-frame exons for GoF or DN variants to downregulate transcript levels, please see "considerations for different inheritance patterns and pathomechanisms" in this section, as well as Section B
- It is theoretically possible to skip an in-frame exon which results in the formation of a stop codon if it is the penultimate exon given that there is some left-over function of the protein (unlikely eligible)
- It is possible to skip adjacent out-of-frame exons if there is a whole exon deletion of an out-of-frame exon to restore the reading frame. Yet, the other criteria on the size of the skipped area (now deletion + skipped exon) and functional domains apply (see below)
- It is theoretically possible to skip consecutive out-of-frame exons so as not to disrupt the reading frame, though designing an ASO that skips two consecutive exons or multiple ASOs may prove to be a challenge
- It is theoretically possible to skip the first coding exon if a nearby in-frame start codon exists, and the first exon meets exon skipping criteria outlined in Table 4

***Assessing Exon Size***

Assessors should consider the size of the exon. As per ClinGen recommendations, an in-frame deletion in the size of 10% or more of the coding transcript is considered a strong criterion for loss of protein function (Walker et al., 2023). However, this is protein and exon-dependent (i.e., losing up to 30% of the dystrophin transcript can still result in a functional, truncated protein (Duan, 2016; Gao & McNally, 2015)). For this reason, skipping an exon that encodes for more than 10% of the protein is considered "unlikely eligible", and obtaining functional evidence is the expected next step.

The percentage of coding region can be calculated as follows:

*Exon size as coding region in % = (exon length in bp/3) / length of the protein in aa *100*

OR

*Exon size as coding region in % = (exon length in bp) / cDNA length of the gene *100*

Please see this example for the NM_003793.4(CTSF):c.264del (p.Cys89fs) variant in video 10. The protein is 484 aa in length. Exon 2 is in-frame 0-0 and spans 99 nucleotides. This information can be obtained from Ensembl, UCSC genome browser, and/or UniProt (see video).

Exon size in coding region % = (99/3) aa / 484 aa * 100 = 6.8 %

There are instances where an exon coding for more than 10% of the protein will be classified as "not eligible". If the exon encodes for more than 10% of the protein and codes for more than one non-repeat domain, this exon should be considered non-skippable and therefore not eligible for exon skipping ASOs. For more detail on assessing the role of functional domains, please see the section titled "Assessing the Role of Functional Domains".

***Searching for Cases of (Natural) Occurrences of Exon Skipping or In-frame Deletions***

Assessors should determine whether exon skipping has already been observed naturally. This information is necessary to further determine whether exon skipping is a suitable option for a given case. There are different ways in which exon skipping can occur naturally and resources such as ClinVar, gnomAD, DECIPHER, ExonSkipDB, and PubMed aid with their identification:

A) Canonical splice site or splice region variants

Assessors should search ClinVar, PubMed, gnomAD, or Decipher to collect data on variants that cause splice aberration leading to full exon skipping. This information should have been validated by sufficient functional data (e.g., RNAseq or qPCR). Note that also variants affecting splicing motifs outside of the splice sites can cause exon skipping and would also fall under this category.

If full exon skipping has been observed and validated and is disease-causing (pathogenic variant), this exon (and therefore variants within that exon) is not eligible for exon skipping therapy. Please note that *in silico* predictions are not to be used as substitutes for RNAseq and qPCR data, and assessors must pay careful attention to what tools were used in the assessment of splicing outcomes of canonical splice site variants.

If full-length exon skipping has been observed in individuals who do not show signs of the disease in question (individuals can of course have other diseases) the exon is eligible for exon skipping. If no evidence of pathogenic or benign exon skipping validated by qPCR or RNAseq exists, assessors should proceed with the analysis.

B) Full exon deletions or in-frame deletions within the exon

Assessors should search ClinVar, PubMed, gnomAD, or Decipher to collect data on in-frame deletions of full-exon deletions within the exon of interest. If in-frame deletions have been observed and validated to be disease-causing (pathogenic), this exon (and therefore variants within that exon) are not eligible. If this is the case, no further analysis is required. However, assessors should pay attention to whether the in-frame deletion creates a stop codon or new amino acid, as these exons are still eligible for exon skipping via ASO (i.e., the pathogenicity is possibly a result of premature termination or the change of an amino acid leading to folding changes and not necessarily the deletion of amino acids itself). If this is the case, assessors should proceed with analysis.

If full exon deletions and in some instances larger in-frame deletions are identified in the general population and classified as benign, this exon can be considered "eligible" for an exon skipping approach.

The assessment for occurrence of natural exon skipping can easily be summarized as follows:

Eligible - Loss of the exon, either due to splice aberration of genomic deletions, have been classified as benign

Not eligible - Loss of the exon or larger parts of the exon have been classified as pathogenic

***Assessing the Role of Functional Domains***

Assessors should also consider the functionality for which the exon codes. To begin, assessors should identify which domains are coded by the exon using tools such as the UCSC Genome Browser. Assessors can then search for the role of these amino acids or domains in the literature using PubMed, or utilize databases such as the Protein Database (PDB) or UniProt. Assessors should pay attention to the role of these specific domains (i.e., DNA binding domain, catalytic domains) or amino acids (i.e., specific amino acid known to play an important role in enzyme activity or protein structure). Sometimes, a web-search with “protein name protein structure” will also yield the desired results.

Though it is difficult to define the importance of the domain or to predict the effects of exon skipping on protein function, assessors can consider the following exons as “not eligible” for exon skipping:

- The exon codes for important amino acid(s) with a known key functional role in the protein, or a functionally validated domain (i.e., involved in a catalytic domain, dimerization domain, inhibitory domain, etc.)
- The exon is a known mutational hotspot for pathogenic LoF missense variants (tools such as MetaDome, Essential3D, and Franklin can be used to help determine this)
- The exon codes for the only functional domain in the protein (i.e., no possibility of residual function remaining)
- The exon codes for multiple functional domains and covers more than 10% of the coding region

An exon that codes for a functional domain that does **not** meet the above criteria means it can be considered as “unlikely eligible”, indicating the need for functional studies.

Additionally, **tandem repeat domains** can also be considered as “unlikely eligible”, assuming the loss of a repeat (or part of a repeat domain) from the protein will still have residual function (Duan, Goemans, Takeda, Mercuri, & Aartsma-Rus, 2021). Assessors can utilize the aforementioned strategies to assess tandem repeat domains. However, in cases where in-frame deletion of a repeat domain have been reported as pathogenic, or have been functionally proven to disrupt protein function, these exons would be considered “not eligible”. Most suitable are in-frame exons that contain a single, full repeat of the tandem repeats and no additional domains. For exon skipping of tandem repeat domains, we recommend that the protein consists of at least 5 tandem repeats.

Generally, we encourage contacting experts on a given gene and protein to discuss the role of the different domains and whether they consider exon skipping a possibility. Further, when looking into functional domains please check the exact role of a domain and the effect of losing the domain. For example, skipping an inhibitory domain could lead to a gain of function effect on protein level.

If the exon codes for less than 10% of the protein, and no functional domain or important amino acids are coded for by the exon and the exon fulfills other criteria listed in Section A, then the exon can be considered as “likely eligible”. Exons are classified using the criteria outlined in Table 4.

**Special considerations for missense variants and small in-frame indels**

If a missense variant or small, in-frame indel lead to a LoF on protein level, additional caution is necessary. These types of variants often indicate that the exon has a specific function even though no domain might be annotated. This could be that the respective amino acids are crucial for folding or the function of a certain domain has not yet been established. In these cases, we recommend paying special attention to other reported pathogenic variants within the exon. Should there be more pathogenic missense variants and in-frame indels than truncating variants, we consider this exon "unlikely eligible" for exon skipping.

However, for missense variants within tandem repeat domains, we refer to the criteria outlined in the tandem repeat domain section as this is an exception to this recommendation.

**Important considerations for different inheritance patterns and pathomechanisms**

The recommendations described above apply without restrictions to LoF variants in recessive disorders. For LoF variants in AD disorders, one might consider the development of allele-specific ASOs. This will ensure that exon skipping does not occur on the wildtype transcript and may result in greater amounts of functional gene product compared to non-allele specific ASOs. Though specific ASO design is beyond the scope of these guidelines, this may prove challenging and limit the types of ASOs to be designed.

In the specific case of an AD disorder with an out-of-frame exon deletion, allele-specificity is necessary otherwise exon skipping is not possible. Here, exon skipping of adjacent out-of-frame exons has the potential to restore the reading frame on the mutant allele, at the same time exon skipping of that out-of-frame exon would destroy the reading frame of the wildtype allele.

Further, for AD disorders associated with LoF variants, an upregulation of protein production from the wildtype allele should also be considered as an alternative option to exon skipping (see Section C).

For GoF and DN variants, additional considerations are possible. The guidelines for exon skipping can be applied as they are, with the exception that toxic GoF caused by the disruption of an inhibitory domain will not be rescued by skipping the exon containing the inhibitory domain (Mohassel et al., 2021). For GoF and DN variants, an out-of-frame exon can be skipped, either the one containing the variant or any other one to downregulate transcript levels. In such a case, haploinsufficiency of the gene should be taken into account which might require allele-selective approaches. Generally, for downregulation of a transcript in case of GoF and DN variants, Section B can be consulted.

**ASO check**

In addition to the recommended assessment strategies, assessors should review the literature or registries for exon skipping ASO strategies. This review can be performed either as the final step to validate your assessment strategy or earlier in the assessment process (see Step 0). Specifically, for exon skipping ASOs, it is crucial to evaluate whether an exon skipping approach has been implemented and validated at both the RNA and protein levels and shown to have the desired effect on the phenotype.

In cases of conflicting evidence of ASO developments and the feasibility of an ASO therapy, consider the quality of the research, the types of experiments conducted, the nature of the results shared, and the publication date. The evaluation of literature on existing ASOs should be carried out at the assessor's discretion, with a critical and discerning approach.

For exon skipping ASOs specifically, the approach will be applicable to all variants within a given exon. However, please pay attention to whether an existing ASO would bind to the variant site and could thus not be used in the case under assessment if it is a different variant. Also check if the existing target site contains a SNP and the same SNP is present on the correct allele in the case under assessment. The exon (and therefore variant) is still considered "eligible" for canonical exon skipping according to the criteria outlined in Table 4 even if the ASO binds to a different variant site. This, however, means that a new ASO needs to be designed and developed for the variant under assessment.

To help with identifying available ASOs for exon skipping, we recommend a search term in Pubmed like this, text in bold would need to be adjusted for the gene and exon being examined:
***ABCA4*** *AND ((ASO) OR (AON) OR (antisense oligonucleotide)) AND (**Exon 17**)*

**Table 4: Classification of variants for their eligibility towards exon skipping.**

| Classification | Criteria |
|---|---|
| Eligible | Evidence that exon skipping does not impair protein function (benign canonical splice site variant leading to exon skipping, benign single exon deletion, naturally occurring transcript does not contain exon, or previously tested ASO with evidence of functional protein product). |
| Likely eligible | Exon is in-frame<br><br>**AND**<br><br>Exon does **NOT** result in ≥10% loss of protein coding sequence if exon is skipped.<br><br>**AND**<br><br>Exon does not create a stop codon when skipped<br><br>**AND**<br><br>Exon does not code for any functional domains<br><br>**AND**<br><br>None of the exclusion criteria outlined in the "Not eligible" section are met. |

| | |
|---|---|
| Unlikely eligible | Exon is in-frame<br><br>**AND**<br><br>Exon does not create a stop codon when skipped<br><br>**AND**<br><br>{<br>Exon results in a loss of ≥10% of coding transcript if exon is skipped **AND/OR** exon codes for a <u>single</u> functional domain<br>}<br><br>**AND**<br><br>None of the exclusion criteria outlined in the “Not eligible” section are met |
| Not eligible[11] | Variant is in an out-of-frame exon **OR**<br>Variant is in first or last coding exon **OR**<br>Variant is in the ONLY coding exon **OR**<br>Exon skipping results in a stop codon **OR**<br>Exon skipping results in the loss of the ONLY functional domain in the protein **OR**<br>Exon encodes for more than 10% of the proteins AND multiple non-repeat domains **OR**<br>Exon codes for functionally proven important domains or amino acids (catalytic site, dimerization domain, inhibitory domains, etc.) **OR**<br>Exon is a known mutational hotspot for (missense) loss-of-function variants **OR**<br>Functional evidence of exon skipping shown to be pathogenic **OR**<br>Functional evidence of in-frame deletions shown to be pathogenic **OR**<br>Evidence that an ASO cannot be developed, shown by two independent investigations at the protein/functional level. Or one investigation with a convincing explanation why an ASO cannot be developed. |

[11] Exceptions apply here for GoF and DN variants. Please see full text.

### Section B - Considerations for Transcript Knockdown

In this section, assessors will be guided on how to assess a variant for eligibility towards knockdown approaches. As described in the Background, knockdown ASOs/siRNAs can bind to the target transcript and downregulate (pre-)mRNA expression. Knockdown strategies can be utilized in cases where the pathomechanism is a result of overexpression, toxic GoF, or DN effects (Lauffer, van Roon-Mom, Aartsma-Rus, & N = 1 Collaborative, 2024). This section covers the following topics:

1. Considerations for pathomechanism
2. Considerations for dosage sensitivity
3. Important considerations for different inheritance patterns and pathomechanisms
4. Strategies for searching the literature for gapmer ASOs and siRNAs

***Considerations for Pathomechanisms***

Variants to be considered for knockdown approaches using gapmer ASOs or siRNA are (toxic) GoF and DN variants and copy number gains. Strategies on how to assess a variant mechanism are discussed in Step 2. Please ensure you have sufficient evidence of GoF or DN pathomechanism before proceeding further.

***Considerations for Dosage Sensitivity***

Before proceeding with the development of a knockdown ASO, it is crucial to consider dosage sensitivity and/or haploinsufficiency. Ideally, a knockdown strategy would be employed when loss-of-function is not expected to cause disease. However, such cases are rare. More commonly, alterations in gene dosage are an underlying cause of disease and must be considered when developing knockdown ASOs. Here it becomes important to be aware of the different inheritance patterns that are implicated in a gene.

If complete loss-of-function is not tolerated, but the loss of one gene copy is, knockdowns can still be considered. Resources such as PubMed, pLI scores, LOEUF scores (both available via gnomAD), DECIPHER dosage sensitivity track (Collins et al., 2022), and ClinGen dosage sensitivity score can be used to determine this. We consider a curation from a reputable, independent source, such as the ClinGen consortium, the highest level of evidence for dosage sensitivity.

An indication whether the loss of one allele is tolerated can also be gained from population databases such as gnomAD; if there are carriers of LoF variants in the general population, it can be assumed that the loss of one allele is safe.. The same considerations apply for carriers of homozygous LoF variants within a gene that implies that the complete loss of this gene is tolerated. Typically, if the loss of one gene copy is not a known cause of disease and has been observed in healthy cohorts, knockdown ASOs can be considered.

Another factor to consider is haploinsufficiency. As described in Step 2, haploinsufficiency refers to a situation in which one healthy, wildtype allele does not generate sufficient protein product to preserve the physiological state (Deutschbauer et al., 2005). One can utilize resources such as gnomAD's pLI and LOEUF scores and ClinGen's dosage sensitivity score (preferred) to determine whether haploinsufficiency is a cause of disease. Typically, a pLI score of equal to or greater than

0.9, the top three deciles of LOEUF scores, or a ClinGen haploinsufficiency score of 3 (sufficient evidence) indicates haploinsufficiency being a cause of disease (please note that this should be verified by functional evidence available in the literature). In cases where haploinsufficiency is a known cause of disease, a knockdown approach should only be considered if the disease caused by haploinsufficiency is less severe than the disorder associated with the GoF or DN variants. Though phenotype considerations are beyond the scope of this guideline, one can consider using OMIM, PubMed, and GeneReviews to assess the genotype-phenotype relation.

One should also consider whether hypomorphic alleles are a cause of disease. Hypomorphic alleles are alleles which show partial loss-of-function (sometimes referred to as “leaky” alleles because there is some retention of protein function). In such cases, it is important to consider ASO dosage and associated phenotypes, especially if partial loss-of-function is disease causing.

**Important considerations for different inheritance patterns and pathomechanisms**

For diseases that are tolerant to complete loss of function (knockout), targeting both alleles may be tolerated. The use of an allele-specific ASO targeting a SNP or the variant site should be considered if changes in gene dosage are a known cause of disease. Though this would greatly limit the ASO design, allele-specific ASOs in these scenarios would help ensure that at least 50% of the wildtype function remains (by targeting only the mutant allele).

Another scenario in which allele-specific ASOs should be considered is if the mechanism is dominant-negative. DN variants impact the wildtype product and therefore may result in a functional product loss of greater than 50%. Hence, it is critical that an ASO is designed to specifically target the DN allele while keeping the wildtype product intact to recapitulate as much function as possible.

In the case of X-linked and Y-linked disorders, further considerations are necessary. For males, downregulation of a gene on the X or Y chromosome can lead to a complete loss of the gene function and before embarking on such an approach it should be known that this complete loss is tolerated. For X-linked disorders in females, it should be considered how X inactivation will affect gene dosage and whether a knockdown approach is safe.

**ASO Check**

In addition to the recommended assessment strategies, assessors should review the literature for knockdown ASO/siRNA strategies. This review can be performed either as the final step to validate the assessment strategy or earlier in the assessment process (see Step 0). Specifically, for knockdown ASOs, it is crucial to evaluate whether a knockdown approach has been developed for other DN or GoF variants in the same gene, and that this approach has been validated at the RNA and protein level and shown to rescue the phenotype (pre-clinical work is sufficient). Take note if an allele-specific approach was used, as this may limit ASO design and affect outcomes. Further, an ASO might have been developed that is specific for a variant other than the one under assessment or a SNP and information on phasing of an individual’s variant with that SNP will then have to be obtained. The gene is still “eligible” for knockdown according to the criteria outlined in Table 5 even if the already available ASO is not suitable for the case under assessment. This means a new ASO would have to be designed and developed for that case.

In cases of conflicting evidence regarding ASO developments and the feasibility of an ASO therapy, consider the quality of the research, the types of experiments conducted, the nature of the results shared, and the publication date. The evaluation of literature on existing ASOs should be carried out at the assessor's discretion, with a critical and discerning approach.

To help with identifying available ASOs/siRNA for knockdown approaches, we recommend a search term in Pubmed like this, text in bold would need to be adjusted for the gene and variant being examined:

***SCN2A*** AND ((ASO) OR (AON) OR (antisense oligonucleotide) OR (AOs) OR (siRNA) OR (RNAi) OR (gapmer) or (knockdown))

***SCN2A*** AND ((ASO) OR (AON) OR (antisense oligonucleotide) OR (AOs) OR (siRNA) OR (RNAi)) AND ((***p.R853Q***) OR (**p.Arg853Gln**) OR (***c.2558G>A***))

Once all this information is collected, one can use Table 5 to classify the variant's eligibility towards ASO knockdowns as "eligible", "likely", "unlikely", or "not eligible". Variants, where not enough evidence exists on the pathomechanism or the dosage sensitivity cannot be assessed until more evidence is collected.

**Table 5: Classification of variant for their eligibility towards knockdown**

| Classification | Criteria |
|---|---|
| Eligible | ASO/RNAi/siRNA has already been developed and shown to work with available functional evidence (i.e., evidence of knockdown rescuing function, pre-clinical data is sufficient) |
| Likely eligible | Variant is Gain-of-Function or Dominant-Negative (functionally proven)<br><br>**AND**<br><br>{<br>Gene is tolerant to the reduction of gene dosage (i.e., gene is NOT haploinsufficient)<br><br>**AND/OR**<br><br>Individuals with heterozygous LoF variants are present in population databases/described in medical literature, such that high penetrance for probably severe disease phenotypes are unlikely<br>} |
| Unlikely eligible | Variant is Gain-of-Function or Dominant-Negative (functionally proven)<br><br>**BUT**<br><br>Heterozygous LoF/Haploinsufficiency/Hypomorphic variants has/have been associated with a disease |

| | |
|---|---|
| Not eligible | Intolerant to reduction (i.e., gene dosage is tightly regulated in humans, and knockdown is expected to lead to serious phenotypic consequences)<br><br>**OR**<br><br>Evidence that an ASO cannot be developed, shown by two independent investigations at the protein/functional level. Or one investigation with a convincing explanation why an ASO cannot be developed. |

### Section C - Considerations for upregulation from the wildtype allele

For disorders caused by haploinsufficiency, one functional wildtype gene copy remains. In these situations, one can use ASOs to upregulate the wildtype allele. This approach, also known as targeted augmentation of nuclear gene output (TANGO), can include skipping of poison exons, downregulating naturally occurring antisense transcripts, and targeting UTR regulator elements such as upstream open reading frames, all of which can increase the gene product. For a more detailed explanation of these approaches, see the Background and the following publications:
1. Lim et al. (2020) https://doi.org/10.1038/s41467-020-17093-9
2. Mittal et al. (2022) https://doi.org/10.1016/j.medj.2022.08.006
3. Felker et al. (2023) https://doi.org/10.1016/j.gim.2023.100884
4. Liu et al. (2022) https://doi.org/10.1093/nar/gkac1094

For convenience, the data found in the supplementary tables and databases from the four aforementioned publications have also been compiled into one excel sheet (Suppl File 4). Assessors can use this sheet to look up poison exons, naturally occurring antisense transcripts, and upstream open reading frames found from each of these publications. It is highly encouraged that assessors do not rely on this one resource alone, but also utilize it in conjunction with the other strategies discussed below.

Please note that unlike the other ASO strategies, these guidelines will not provide details on how to classify variants as “likely”, “unlikely", or “not eligible”. The upregulation of wildtype transcripts through the aforementioned strategies heavily depends on the availability of functional evidence for key regulatory elements. These strategies are not comparable to one another, and different approaches must be employed depending on the availability of regulatory elements. Instead, readers can reference this section to learn about the different strategies and resources they can utilize for their own analyses. However, in case the case of a wildtype upregulation approach has already been established for a given gene with sufficient functional evidence, we consider a variant as “eligible” toward wildtype upregulation. This section covers:

1. Targeting naturally occurring antisense transcripts
2. Targeting upstream open reading frames
3. Targeting poison exons and non-productive alternative splicing events

**Naturally Occurring Antisense Transcripts**

Through downregulating naturally occurring antisense transcripts via gapmer ASOs, it is possible to upregulate gene expression (Fig. 3). Assessors can utilize resources such as PubMed, UCSC Genome Browser, HUGO Gene Nomenclature Committee, and Ensembl to search for known antisense transcripts for a given gene. Additionally, the supplemental table 2 in the paper by Mittal et al. (2022) has listed all antisense transcripts the authors have identified. Assessors should also consider the level at which these antisense transcripts are expressed in the tissue of interest. Databases such as GTEx and the Human Protein Atlas can assist with this. Note that the existence of an antisense transcripts alone is not enough to consider an ASO development. Factors such as tissue expression, regulatory function, and orientations of the antisense transcripts must be well understood before considering this approach.

**Upstream Open Reading Frames**

An ASO can be designed to targets the regulatory elements in untranslated regions, including uORFs (Fig. 3). Resources such as PubMed and Ribo-uORF can be used to check whether there is a uORF that can be targeted. Additionally, the supplemental table 2 in the paper by Mittal et al. (2022) indicates for which genes uORFs have been identified. Note that not all uORFs act through an inhibitory mechanism, and proper characterization of their mechanism is essential in determining their eligibility as ASO targets. Another possible approach is skipping an exon in the UTR that contains a uORF.

**Poison Exons and Non-Productive Alternate Splicing Events**

One can design ASOs which target non-productive alternate splicing events including poison exons (Fig. 3). These ASOs would utilize the splice-switching approaches described in the splice-correction and exon skipping sections to promote canonical splicing of the wildtype transcript. Resources such as PubMed, Ensembl, VastDB, and the UCSC Genome Browser can aid in determining alternate splicing events in the transcript of interest. Additionally, supplemental table 2 from Mittal et al. (2022), supplemental data 2 from Lim et al. (2020), and supplemental data 1 from Felker et al. (2023) lists all identified poison exons in these papers. Databases such as GTEx can further assist by determining the expression level of alternate transcripts in the target tissue. Note that some alternate splicing events are crucial for the production of important transcripts and isoforms.

For more resources and strategies on upregulating wildtype gene product, please reference the *Useful Tools* section.

#### Important considerations for different inheritance patterns and pathomechanisms

The considerations outlined here are mainly applicable to diseases associated with haploinsufficiency, which is most likely caused by LoF variants in AD disorders. In rare cases, one could also consider applying upregulation of wildtype allele strategies in X-linked disorders in females where there is sufficient evidence that upregulation from the second X chromosome is possible. Please always consider challenges caused due to X inactivation.

#### ASO Check

Also for the upregulation of wildtype allele approaches, one can search whether specific strategies already exist, are under development or pursued in clinical trials. Multiple strategies can be utilized to upregulate gene product from wildtype transcripts, and it is therefore important to employ a variety of strategies/approaches in the search terms to ensure a comprehensive review of the field.

While we do not classify the different upregulation approaches, we consider a case/variant as “eligible” for upregulation if an upregulation strategy has been developed and demonstrated to work with sufficient evidence.

### Addendum

**Summary "Unable to assess"**

A variant is classified "unable to assess" when:

- The variant does not fall into the category of variants for these guidelines, e.g. translocations, variants in non-coding genes
- The variant description is incorrect
- The inheritance pattern is not known
- The pathomechanism of the variant is not known
- Not enough information on dosage sensitivity available (for knockdown)
- Loss of both copies of a gene due to large/whole gene deletions
- Intronic variant without sufficient functional information on splicing

### Examples

The below examples provide variant assessments for different types of variants. Detailed explanations for all the examples can be found in the accompanying training videos (via YouTube or the N1C website).

To further practice variant assessments, we encourage new assessors to use the test variants in Suppl File 2 and later check the answer keys in Suppl File 3.

**Table 6: Example variants and their assessments**

| # | Variant and Video Content | Assessment |
|---|---|---|
| 1 | NM_000350.3(ABCA4):c.2626C>T p.(Gln876Ter)<br><br>Corresponding video content:<br>• Nomenclature check using Mutalyzer<br>• Searching UCSC to determine exon number<br>• Searching PubMed for exon skipping ASO<br>• Recommendations for continued analysis | **Eligibility: Eligible for exon skipping ASO**<br><br>Explanation: Correct variant description, variant is loss of function in a recessive disorder. Variant is >15 bp downstream of the nearest splice site and is a nonsense variant considered for exon skipping.<br><br>Variant is located in exon 17/50 in the *ABCA4* gene, exon is a small, in-frame exon. Skipping of in-frame exon 17 has been shown in pre-clinical studies (Kaltak et al., 2023). |
| 2 | NM_016589.4(TIMMDC1):c.597-1340A>G<br><br>Corresponding video content:<br>• Nomenclature check using VariantValidator (and example output of incorrect variant) | **Eligibility: Eligible for splice correction ASO**<br><br>Explanation: Correct variant description, variant is loss of function in a recessive gene. Variant is deep intronic and >100 bp from the nearest splice site. Variant has been reported multiple times and the effect on splicing has |

| | | |
|---|---|---|
| | ● Searching PubMed for splice-switching ASO | been confirmed via RNAseq (Kremer et al., 2017).<br><br>Variant causes the insertion of a cryptic exon and a premature stop.<br><br>Pre-clinical data is available that shows evidence of rescue of enzymatic effects upon ASO treatment (Kumar et al., 2022). |
| 3 | NM_000533.5(PLP1):c.680dup p.(Cys228LeufsTer5)<br><br>Corresponding video content:<br>● Checking inheritance patterns using OMIM<br>● Checking variant mechanism using GeneReviews<br>● Using UCSC genome browser to determine exon position<br>● Using ExonViz to determine exon frame | **Eligibility: Not eligible for exon skipping ASO**<br><br>Explanation: Correct variant description, X-linked recessive gene, frameshift variant leading to an early stop can only be considered for exon skipping (Section A). Variant >15 bp upstream of nearest splice site. Exon 5/7 is out-of-frame and thus not eligible. |
| 4 | NM_003793.4(CTSF):c.213+1G>C<br><br>Corresponding video content:<br>● Checking inheritance patterns using OMIM<br>● Checking variant mechanism using ClinVar<br>● Assessing splicing effects of variants through a literature search | **Eligibility: Not eligible for splice correction ASO**<br><br>Explanation: Correct variant description, autosomal recessive gene. Variant is a canonical splice site variant and was shown to lead to skipping of exon 1, leading to disease (Di Fabio et al., 2014). Variant is within 5 bp of canonical splice site. |
| 5 | NM_000277.3(PAH):c.611A>G p.(Tyr204Cys)<br><br>Corresponding video content:<br>● Checking inheritance patterns using OMIM<br>● Checking variant mechanisms using OMIM and a literature search<br>● Assessing splicing effects of variants through a literature search<br>● Assessing effects of missense variant through a literature search<br>● Checking the position of the variant respective to canonical splice sites using UCSC | **Eligibility: Unlikely for splice correction ASO**<br><br>Explanation: Correct variant description, variant is in a recessive gene, variant is a missense variant with evidence of effect on splicing.<br><br>Variant affects mRNA splicing and results in a 32-amino acid deletion of the PAH enzyme (Ellingsen, Knappskog, & Eiken, 1997). Variant is classified as unlikely since Ellingsen et al. could only identify a small change in enzymatic activity when generating the enzyme with the missense variant. |

| | | |
|---|---|---|
| 6 | NM_025152.3(NUBPL):c.815-27T>C<br><br>Corresponding video content:<br>● Checking inheritance patterns using OMIM<br>● Assessing variant mechanisms and splicing effects through a literature search | **Eligibility: Unlikely eligible for splice correction ASO**<br><br>Explanation: Correct variant description. Variant associated with autosomal recessive disorder. Variant is intronic with evidence of effect on splicing.<br><br>Variant is not within 5bp of the canonical splice site, but in the 5 to 100 bp region. Variant affects a branchpoint. The branchpoint is weakened but not destroyed (30% of wildtype transcript remains) (Maclean, Kimonis, & Balk, 2018). Therefore, this variant is unlikely eligible for splice correction ASO. |
| 7 | NM_024312.5(GNPTAB):c.3503_3504del p.(Leu1168fs)<br><br>Corresponding video content:<br>● Searching the literature for an already existing ASO<br>● Checking inheritance patterns using OMIM<br>● Assessing variant mechanisms and eligibility for exon-skipping using the literature | **Eligibility: Unlikely eligible for exon skipping ASO**<br><br>Explanation: Correct variant description. ASO exists but has only been validated at the RNA level (not the protein level). Variant is associated with autosomal recessive disorder and is loss-of-function.<br><br>Variant is in exon 19/21. Exon is in-frame and codes for 4.5% of the coding transcript. A stop codon does not form when exon is skipped. Exon codes for a stealth domain, but does not meet any of the exclusion criteria in Table 3 (Matos et al., 2020). Therefore, this variant is unlikely eligible for an exon skipping ASO. |
| 8 | NM_001040142.2(SCN2A):c.5645G>A p.(R1882Q)<br><br>Corresponding video content:<br><br>● Nomenclature check using variant validator<br>● Inheritance pattern check using OMIM<br>● Pathomechanism check using ClinVar and by conducting a literature search<br>● Dosage sensitivity assessment using ClinGen, pLI, and the literature<br>● Conducting an ASO check | **Eligibility: Eligible for knockdown**<br><br>Explanation: Correct variant description. ASO exists and has been validated clinically. Variant is associated with an autosomal dominant gain-of-function disorder.<br><br>Haploinsufficiency and loss-of-function is also a known cause of disease, but is arguably associated with less severe phenotypes. Variant classified as “eligible” because preclinical evidence of knockdown approach exists. |

| | | |
|---|---|---|
| 9 | NM_001165963.4(SCN1A):c.3733C>T p.(R1245Ter)<br><br>Corresponding video content:<br>● Nomenclature check using variant validator<br>● Inheritance pattern check using OMIM<br>● Pathomechanism check using ClinVar<br>● Dosage sensitivity assessment using ClinGen and pLI<br>● Checking for upregulation (i.e. TANGO) methods using the literature<br>● Conducting an ASO check<br>● Brief discussion on other upregulation of wildtype allele approaches using NM_130839.5 (UBE3A):c.67C>T, p.(Arg23*) as an example | **Eligibility: Eligible for upregulation of wildtype allele**<br><br>Explanation: Correct variant description. ASO exists and has been clinically validated. Variant is associated with autosomal dominant loss-of-function disorder. Haploinsufficiency and loss-of-function is a known cause of disease.<br><br>Variant is found in an exon not eligible for canonical exon skipping.<br><br>Poison exons identified in the literature, and have been clinically validated. |
| 10 | NM_003793.4(CTSF):c.264del p.(Cys89fs)<br><br>Corresponding video content:<br>● Checking inheritance patterns using OMIM<br>● Checking variant mechanism using ClinVar<br>● Using UCSC genome browser to determine exon position<br>● Using ExonViz and Ensembl to determine exon frame<br>● Using RefSeq and UniProt to determine exon size<br>● Using UniProt to assess corresponding protein function<br>● Recommendations for continued analysis | **Eligibility: Likely eligible for exon skipping ASO**<br><br>Explanation: Correct variant description, autosomal recessive gene, variant is a frameshift variant, thus can only be assessed for exon skipping (Section A). Exon 2/13 is in-frame, approx. 7% of the protein coding region. Skipping of exon does not create a stop codon. No functional domain is known and no mutational hotspot identified. |
| 11 | NM_000170.3(GLDC):c.538C>T p.(Gln285Ter)<br><br>Corresponding video content:<br>● Inheritance pattern check using OMIM | **Eligibility: Not eligible for exon skipping**<br><br>Explanation: Variant description is correct. Variant is associated with autosomal recessive loss-of-function disorder. Variant is not in the first or last codon exon. Variant is not in an out- |

| | | |
|---|---|---|
| | • Pathomechanism check using ClinVar<br>• Using UCSC genome browser to determine exon position and frame<br>• Using UCSC to check whether exon skipping forms a stop codon | of-frame exon. However, variant is found in an exon which will form a stop codon if skipped, and the two neighboring exons join together. Therefore, this variant is not eligible towards canonical exon skipping ASOs. |
| 12 | Unable to Assess Examples<br><br>Corresponding video content:<br><br>• Example of a mitochondrial variant<br>• Example of incorrect variant notation<br>• Example of a missense variant with an unknown pathomechanism | **Eligibility: Unable to assess**<br><br>All variants discussed in the video are ineligible for assessment either because they are unable to be assessed by these specific guidelines, or not enough information is available to proceed with assessment. |

### Useful Tools

This is a collection of tools that can be useful during variant assessment. Most of them are already listed throughout the text. Tools are listed matching the different steps of these guidelines.

**Step 0 - Variant check**
HGVS nomenclature:https://varnomen.hgvs.org/
Mutalyzer: https://mutalyzer.nl/ (cannot do deep intronic variants in c. notation but works with g. notation)
VariantValidator: https://variantvalidator.org/

**Step 1 - Assessment of pattern of inheritance and disease type**
Gene Cards: http://www.genecards.org/
OMIM: https://omim.org/
Gene Reviews: https://www.ncbi.nlm.nih.gov/books/NBK1116/
Orphanet: https://www.orpha.net/en/disease
Monarch Initiative: https://monarchinitiative.org
Gene Curation Coalition: https://thegencc.org/
ClinGen: https://www.clinicalgenome.org/
gnomAD: https://gnomad.broadinstitute.org/ (population frequency of a variant)
DisGeNet: https://www.disgenet.org/
Gene2Phenotype: https://www.ebi.ac.uk/gene2phenotype

**Step 2 - Assessment of pathomechanism of genetic variant**
HGMD: https://www.hgmd.cf.ac.uk/
DECIPHER: www.deciphergenomics.org/
ClinVar: https://www.ncbi.nlm.nih.gov/clinvar/

Mastermind Genomenon: https://mastermind.genomenon.com/
Pubmed: https://pubmed.ncbi.nlm.nih.gov/
Franklin: https://franklin.genoox.com/clinical-db/home
LOVD: https://www.lovd.nl/
Varsome: https://varsome.com/

**Step 3 - Evaluation of splicing effects**
HGMD: https://www.hgmd.cf.ac.uk/
DECIPHER: https://www.deciphergenomics.org/
ClinVar: https://www.ncbi.nlm.nih.gov/clinvar/
Mastermind Genomenon: https://mastermind.genomenon.com/
Pubmed: https://pubmed.ncbi.nlm.nih.gov/
Search enginer → web-search the variant

**Section A - Considerations for canonical exon skipping**
UCSC Genome Browser:https://genome.ucsc.edu/
ExonViz:https://exonviz.rnatherapy.nl/
Ensembl: https://www.ensembl.org/index.html
Metadome: https://stuart.radboudumc.nl/metadome/
UniProt: https://www.uniprot.org/
Protein Database: https://www.rcsb.org/
Alphafold: https://alphafold.ebi.ac.uk/
ES-NDD: https://es-ndd.broadinstitute.org/
Franklin: https://franklin.genoox.com/clinical-db/home
ExonSkip DB: https://ccsm.uth.edu/ExonSkipDB/

**Section B - Considerations for Downregulation**
gnomAD for pLI and Loef: https://gnomad.broadinstitute.org/
Dosage sensitivity map:UCSC Genome Browser: News Archives
ClinGen dosage sensitivity curation: https://search.clinicalgenome.org/kb/gene-dosage?page=1&size=25&search=
DECIPHER dosage sensitivity track: https://genome.ucsc.edu/goldenPath/newsarch.html#022124

**Section C - Considerations for upregulation from the wildtype allele**
Antisense transcripts via HUGO: Search results | HUGO Gene Nomenclature Committee (genenames.org)
Ribo uORF:https://rnainformatics.org.cn/RiboUORF/
GTEx: https://gtexportal.org/home/transcriptPage
VastDB: https://vastdb.crg.eu/wiki/Main_Page

**Further tools**
MobiDetails: https://mobidetails.iurc.montp.inserm.fr/MD/
ProteinPaint: https://proteinpaint.stjude.org/
Proteinatlas:http://www.proteinatlas.org/
FLIBase - Full length isoforms in cancers and normal tissues: http://flibase.org/#/home

### Abbreviations

AD autosomal dominant

AR autosomal recessive

ASO Antisense Oligonucleotide

CNV company number variant

DN dominant negative

GoF gain of function

HGVS Human Genome Variation Society

LoF loss of function

NMD nonsense mediated decay

ORF open reading frame

pORF primary open reading frame

SE splice enhancer

siRNA small interfering RNA

SNP Single nucleotide polymorphism

SS splice silencer

TANGO targeted augmentation of nuclear gene output

uORF upstream open reading frame

UTR untranslated region

WT wildtype
