## Supplemental File 2 for "Consensus guidelines for eligibility assessment of pathogenic variants to antisense oligonucleotide treatments"

**Supplementary File 2**

Table with test variants for piloting rounds. The table indicates which test variants were provided to the assessors for the 3 piloting rounds. This purposely includes variants with incorrect descriptions.

| Piloting Round | Transcript | Gene | Variant |
|---|---|---|---|
| **1** | NM_025152.3 | *NUBPL* | c.815-27T>C |
| **1** | NM_000350.3 | *ABCA4* | c.769-784C>T |
| **1*** | NM_024312.5 | *GNPTAB* | c.3505_3504del |
| **2** | NM_020366.3 | *RPGRIP1* | c.1468-128T>G |
| **2** | NM_000329.3 | *RPE65* | c.1430A>G |
| **2** | NM_001086521.2 | *NDUFAF8* | c.195+271C>T |
| **2** | NM_003907.3 | *EIF2B5* | c.1156+13G>A |
| **2** | NM_206933.4 | *USH2A* | c.2692C>T |
| **2** | NM_000391.4 | *TPP1* | c.225A>G |
| **2** | NM_024298.5 | *MBOAT7* | c.758_778del |
| **2** | NM_018075.5 | *ANO10* | c.289del |
| **2*** | NM_024312.5 | *GNPTAB* | c.3503_3504del |
| **2** | NM_003650.4 | *CST7* | c.2035-946G>A |
| **2** | NM_000303.3 | *PMM2* | c.640-15479C>T |
| **2** | NM_000202.8 | *IDS* | c.1122C>T |
| **3** | NM_018075.5 | *ANO10* | c.1025G>A |
| **3** | NM_001127222.2 | *CACNA1A* | c.4174G>A |
| **3** | NM_024312.5 | *GNPTAB* | c.3488del |
| **3** | NM_133433.4 | *NIPBL* | c.5329-15A>G |
| **3** | ENST00000361390.2 | *MT-ND1* | m.4142G>T |
| **3** | NM_014727.3 | *KMT2B* | c.8079delC |
| **3** | NM_024312.5 | *GNPTAB* | c.1123C>T |
| **3** | NM_005859.5 | *PURA* | c.159dup |
| **3** | NM_001167623.2 | *CACNA1C* | c.1216G>A |
| **3** | NM_000561.4 | *HEXB* | c.1509-26G>A |
| **3** | ENST00000435607.3 | *SCN4A* | c.3891C>A |
| **3** | NM_001244008.2 | *KIF1A* | c.914C>T |
| **3** | NM_000492.4 | *CFTR* | c.2989-313A>T |
| **3** | NM_001194.4 | *HCN2* | c.736G>A |
| **3** | NM_000170.3 | *GLDC* | c.538C>T |

*Indicates duplicate variants assessed in multiple rounds
