## Supplemental File 3 for "Consensus guidelines for eligibility assessment of pathogenic variants to antisense oligonucleotide treatments"

**Supplementary File 3**

Answer keys to all test variants assessed in the 3 piloting rounds.
Classification of all variants was corrected for the classifications in accordance with the latest version of the guidelines (v1.0 September 2024).

### ROUND 1

**Variant 1: *NUBPL* - NM_025152.3:c.815-27T>C**

- Variant description is correct, no ASO exists.
- Associated disorder is autosomal recessive (https://www.omim.org/entry/613621)
- Variant is loss-of-function
- Variant affects a branchpoint and leads to aberrant splicing (https://doi.org/10.1093/hmg/ddy247 and https://doi.org/10.1002/humu.21654)
- Branchpoint is not fully destroyed (30% of the wildtype transcript remains)
- Variant is intronic and not within 15 base pairs of canonical splice site, but branchpoint is weakened.
- **Therefore, the variant is unlikely eligible for splice correction**

**Variant 2: *ABCA4* - NM_000350.3:c.769-784C>T**

- Variant description is correct, ASO exists (10.3390/cells11243947), as proven through studies on patient-derived fibroblasts
- **Therefore, this variant is classified as eligible for splice correction**

**Variant 3: *GNPTAB* - NM_024312.5:c.3503_3504del**

- Variant description is correct, ASO has been studied (https://doi.org/10.1089/hum.2020.034), however, this paper did not validate the effect of exon-19 skipping on protein function. In their discussion, the authors noted that the effect of this ASO on protein expression, subcellular localization, cleavage of the GlcNAc-1-phosphotransferase, and correction of mis-sorting of lysosomal enzymes needs to be studied.
- Additionally, exon 19 codes for a repeat stealth domain. However, this domain does not meet the criteria for exclusion (i.e., it is not the only domain, it is not a mutation hotspot, and it is not functionally proven to be important).
- The following was noted in the discussion:
    - *"However, unlike the Stealth domains 1-3 harboring a high number of ML II-causing missense mutations, only one ML III alpha/beta causing amino-acidic substitution has been reported on the fourth Stealth domain [c.3458A>G; p.(Asn1153Ser)]. Furthermore, a recent combined in vitro and in silico analysis of missense GNPTAB mutations has provided new insights into the role of these conserved Stealth regions for catalytic activity of GlcNAc-1-phosphotransferase, showing that the amino acid residues Glu389, Asp408, His956, and Arg986 are strictly required for enzymatic catalysis. Interestingly, none of these residues locates on the fourth Stealth domain, suggesting that it is less important for GlcNAc-1-phosphotransferase activity."*
- **Therefore, the variant is unlikely eligible for exon skipping**

| ROUND 2 |
| --- |

**Variant 1: *RPGRIP1*- NM_020366.3:c.1468-128T>G**
- Variant description is correct
- ASO exists (10.3390/genes12020287)
- **Therefore, this variant is classified as <u>eligible</u> for splice correction**

**Variant 2: *RPE65* - NM_000329.3:c.1430A>G**
- Variant description is correct
- No ASO exists
- Variant is inherited in an autosomal dominant manner and variant is dominant negative (10.3390/genes11121420)
- During round 2 of the piloting, the variant was classified as "<u>unable to assess</u>" since this round did not include autosomal dominant inheritance and dominant negative variants. However, with the extended guidelines, this variant can now be assessed.
- If assessed using the current guidelines, which are also applicable to dominant negative variants, the variant would be **<u>likely eligible</u> for both splice correction and knockdown**.
- Splice correction: The variant and novel acceptor site is not within 6 to 15 basepairs of the canonical splice site, and forced expression of the D477G protein behaved like wildtype RPE65 protein (10.1002/humu.23706).
- Knockdown: Variant is dominant negative, and haploinsufficiency is not a known cause of disease (pLI score of 0, ClinGen haploinsufficiency score is "autosomal recessive").

**Variant 3: *NDUFAF8* - NM_001086521.2:c.195+271C>T**
- Variant description is correct
- No ASO exists
- Variant is inherited in an autosomal recessive manner and variant is loss-of-function variant
- Functional evidence of aberrant splicing, determined using cDNA studies on RNA derived from patient fibroblasts (https://doi.org/10.1016/j.ajhg.2019.12.001)
    - Traces of wildtype transcript can be seen in the supplementary figures.
    - The variant meets all criteria to be established as a "probably" (intronic, not within 15 base pairs).
    - Note: It's important to make your own judgement using available data. The article stated degradation of the transcript is associated with this variant, but trace amounts can be seen in the supplementary. Furthermore, consider the location of the variant and whether its position can possibly destroy a branchpoint (typically found within 40-80bp of the 3' of the intron) or canonical splice site.
- **Therefore, this variant is <u>likely</u> eligible for splice correction**

**Variant 4: *EIF2B5* - NM_003907.3:c.1156+13G>A**
- Variant description is correct

- No ASO exists
- Variant is inherited in an autosomal recessive manner and variant is loss-of-function variant
- Functional evidence of aberrant splicing, determined using cDNA studies on RNA derived from patient cells (https://www.ncbi.nlm.nih.gov/pmc/articles/PMC7480926/)
    - Minigene assays show that 20% of the wildtype transcript is still expressed (note that we advise against the use of minigene assays as sufficient evidence in our guidelines)
- Variant is within +50 base pairs of canonical splice site
- **Therefore, this variant is <u>unlikely</u> eligible for splice correction**

**Variant 5: *USH2A* - NM_206933.4:c.2692C>T**
- Variant description is correct
- ASO exists with functional evidence at the protein level (https://www.cell.com/molecular-therapy-family/molecular-therapy/fulltext/S1525-0016(21)00212-4)
- **Therefore, this variant is classified as <u>eligible</u> for canonical exon skipping**

**Variant 6: *TPP1* - NM_000391.4:c.225A>G**
- Variant description is correct
- No ASO exists
- Variant is inherited in an autosomal recessive manner and variant is a loss-of-function variant
- Functional evidence of aberrant splicing determined through RNAseq (https://doi.org/10.1016/j.ejmg.2021.104259)
- Variant is within 5 base pairs of the canonical splice site
- **Therefore, this variant is <u>not eligible</u> for splice correction**

**Variant 7: *MBOAT7* - NM_024298.5:c.758_778del**
- Variant description is correct
- No ASO exists
- Variant is inherited in an autosomal recessive manner
- Functional evidence for this specific variant does not exist
- **Therefore, this variant is classified as "<u>unable to assess</u>"**
- **Note**: Even if functional evidence existed and this variant was classified as loss-of-function, this variant results in an in-frame deletion. Pathogenic variants resulting from in-frame deletions may suggest that the pathomechanism is a result of the deletion itself, and that deleting the entire exon (even if in-frame) will not ameliorate the pathogenicity.

**Variant 8: *ANO10* - NM_018075.5:c.289del**
- Variant description is correct
- No ASO exists
- Variant is inherited in an autosomal recessive manner and the variant is a loss-of-function variant.

- Variant is found in exon 3:
    - Exon is in-frame (198 base pairs)
    - Exon does not code for a stop codon if skipped
    - Exon codes for EXACTLY 10% of the coding transcript (66 amino acids coded for by exon 3, total protein length is 660 amino acids).
    - Exon codes for a cytoplasmic domain (though the role of this domain is not functionally proven or considered essential to protein function, to our knowledge)
- **Therefore, this variant is classified as <u>unlikely</u> eligible for exon skipping**

**Variant 9: *GNPTAB* - NM_024312.5:c.3503_3504del**

- Variant is correct, ASO has been studied (https://doi.org/10.1089/hum.2020.034), however, this paper did <u>not</u> validate the effect of exon-19 skipping on protein function. In their discussion, the authors noted that the effect of this ASO on protein expression, subcellular localization, cleavage of the GlcNAc-1-phosphotransferase, and correction of mis-sorting of lysosomal enzymes needs to be studied.
- Additionally, exon 19 codes for a repeat stealth domain. However, this domain does <u>not</u> meet the criteria for exclusion (i.e. it is not the <u>only</u> domain, it is not a mutation hotspot, and is not functionally proven to be important).
- The following was noted in the discussion:
    - *"However, unlike the Stealth domains 1-3 harboring a high number of ML II-causing missense mutations, only one ML III alpha/beta causing amino-acidic substitution has been reported on the fourth Stealth domain [c.3458A>G; p.(Asn1153Ser)]. Furthermore, a recent combined in vitro and in silico analysis of missense GNPTAB mutations has provided new insights into the role of these conserved Stealth regions for catalytic activity of GlcNAc-1-phosphotransferase, showing that the amino acid residues Glu389, Asp408, His956, and Arg986 are strictly required for enzymatic catalysis. Interestingly, none of these residues locates on the fourth Stealth domain, suggesting that it is less important for GlcNAc-1-phosphotransferase activity."*
- **Therefore, the variant is <u>unlikely</u> eligible for canonical exon skipping**

**Variant 10: *CST7* - NM_003650.4:c.2035-946G>A**

- Variant description is incorrect
- **Therefore, this variant is classified as "<u>unable to assess</u>"**

**Variant 11: *PMM2* - NM_000303.3:c.640-15479C>T**

- Variant description is correct
- ASO exists with functional evidence at the protein level (https://pubmed.ncbi.nlm.nih.gov/19235233/)
- From the abstract by Chen *et al*. (2024): "*Here we developed antisense oligonucleotides (ASOs) to effectively decrease the inclusion of exon 8A in human cells both in vitro and, following transplantation, in vivo. We discovered*

*that the ASO-mediated switch from exon 8A to 8 robustly rescued defects in patient-derived cortical organoids and migration in forebrain assembloids"*

- **Therefore, this variant is classified as eligible for splice correction**

**Variant 12: *IDS* - NM_000202.8:c.1122C>T**

- Variant description is correct
- ASOs were studied but were unsuccessful (https://pubmed.ncbi.nlm.nih.gov/26407519/). However, unsuccessful splice-correcting ASOs are not exclusion criteria (this is only applicable for exon skipping ASOs that show exon skipping to be pathogenic). Variant is autosomal recessive and loss-of-function
- Functional evidence of splicing
- Variant is synonymous, not within 15 base pairs of the canonical splice site, and is not thought to weaken or destroy the canonical splice sites.
- **Therefore, this variant is likely eligible for splice correction**
- *Note: If multiple groups show unsuccessful development or there is evidence why an ASO will not work, we do consider the variant as not eligible.*

### ROUND 3

**Variant 1: *ANO10* - NM_018075.5:c.1025G>A**

- Variant description is correct, no ASO exists.
- Variant is inherited in an autosomal recessive manner and a loss-of-function variant, and therefore we can consider canonical exon skipping.
- Exon codes for 28% of transcript and a transmembrane domain.
- **Larger exons which code for multiple domains are considered as not eligible. Therefore, this variant should be classified as not eligible towards canonical exon skipping ASOs.**

**Variant 2: *CACNA1A* - NM_001127222.2:c.4174G>A**

- Variant description is correct, no ASO exists.
- Variant is inherited in an autosomal dominant manner and a gain-of-function variant. Variant is associated with severe developmental and epileptic encephalopathy. However, haploinsufficiency is also a known cause of disease (https://pubmed.ncbi.nlm.nih.gov/31468518/ and https://search.clinicalgenome.org/kb/conditions/MONDO:0007163).
- **Therefore, variant is unlikely eligible for knockdown**.

**Variant 3: *GNPTAB* - NM_024312.5:c.3488del**

- Variant description is correct, exon skipping ASO has been studied for this exon (https://doi.org/10.1089/hum.2020.034). However, this paper did not validate the effect of exon-19 skipping on protein function. In their discussion, the authors noted that the effect of this ASO on protein expression, subcellular localization, cleavage of the GlcNAc-1-phosphotransferase, and correction of mis-sorting of lysosomal enzymes needs to be studied.

- Additionally, exon 19 codes for a repeat stealth domain. However, this domain does not meet the criteria for exclusion (i.e it is not the only domain, it is not a mutation hotspot, and is not functionally proven to be important).
- The following was noted in the discussion:
    - *"However, unlike the Stealth domains 1-3 harboring a high number of ML II-causing missense mutations, only one ML III alpha/beta causing amino-acidic substitution has been reported on the fourth Stealth domain [c.3458A>G; p.(Asn1153Ser)]. Furthermore, a recent combined in vitro and in silico analysis of missense GNPTAB mutations has provided new insights into the role of these conserved Stealth regions for catalytic activity of GlcNAc-1-phosphotransferase, showing that the amino acid residues Glu389, Asp408, His956, and Arg986 are strictly required for enzymatic catalysis. Interestingly, none of these residues locates on the fourth Stealth domain, suggesting that it is less important for GlcNAc-1-phosphotransferase activity."*
- **Therefore, the variant is <u>unlikely</u> eligible for canonical exon skipping.**

**Variant 4: *NIPBL* - NM_133433.4:c.5329-15A>G**
- Variant description is correct, no ASO exists.
- Variant is inherited in an autosomal dominant manner.
- Variant results in alternate splicing. Variant is thought to affect a branchpoint and leads to skipping of in-frame exon 28. Both wildtype and alternately spliced transcripts were detected in patient (https://www.ncbi.nlm.nih.gov/pmc/articles/PMC4746300/).
- **Therefore, the variant is <u>unlikely</u> eligible for splice correcting ASO.**
- **Variant can also be considered for upregulation from the wildtype allele.**

**Variant 5: MT-ND1 - ENST00000361390.2:m.4142G>T**
- Variant is mitochondrial and therefore non-eligible for assessment with these guidelines.
- **Therefore, we are <u>unable to assess</u> this variant.**

**Variant 6: *KMT2B* - NM_014727.3:c.8079delC**
- Variant description is correct, no ASO exists.
- Variant is inherited in an autosomal dominant manner.
- Cannot determine or predict pathomechanism. Variant results in frameshift in the last exon, and therefore cannot assume loss-of-function or nonsense mediated decay. Gain-of-function mechanisms have also been reported as a cause of disease.
- To our current knowledge, no loss-of-function variant exists downstream of this variant.
- **Therefore, this variant is classified as <u>unable to assess</u>.**

**Variant 7: *GNPTAB* - NM_024312.5:c.1123C>T**
- Variant description is correct, no ASO exists.

- Variant is inherited in an autosomal recessive manner and variant is loss-of-function (NP_077288.2:p.(Arg375Ter)).
- Variant can be assessed for canonical exon skipping.
- Variant is in exon 10, which contains functionally important residues (https://doi.org/10.1089/hum.2020.034), and has been reported in a patient with exon 10 skipping (https://www.ncbi.nlm.nih.gov/pmc/articles/PMC6800466/).
- **Therefore, this variant is not eligible for canonical exon skipping.**

**Variant 8: *PURA* - NM_005859.5:c.159dup**

- Variant description is correct, no ASO exists.
- Variant is inherited in an autosomal dominant manner and variant is loss-of-function.
- The variant can be assessed for canonical exon skipping, however, it is a single exon gene.
- **Therefore, this variant is not eligible for canonical exon skipping.**
- **The variant can also be considered for upregulation from the WT allele.**

**Variant 9: *CACNA1C* - NM_001167623.2:c.1216G>A**

- Variant description is correct.
- Variant is inherited in an autosomal dominant manner and variant is gain-of-function.
- ASO exists, and has been validated at the protein level: https://www.nature.com/articles/s41586-024-07310-6
- From the abstract by Chen *et al*. (2024): "*Here we developed antisense oligonucleotides (ASOs) to effectively decrease the inclusion of exon 8A in human cells both in vitro and, following transplantation, in vivo. We discovered that the ASO-mediated switch from exon 8A to 8 robustly rescued defects in patient-derived cortical organoids and migration in forebrain assembloids*"
- **Therefore, this variant is considered eligible towards canonical exon skipping.**

**Variant 10: *HEXB* - NM_000561.4:c.1509-26G>A**

- Variant description is incorrect.
- **Therefore, we are unable to assess this variant.**

**Variant 11: *SCN4A* - ENST00000435607.3:c.3891C>A**

- Variant description is correct, no ASO exists.
- No functional evidence exists for this variant. Variant is assumed to be gain-of-function, but has not been functionally proven: https://doi.org/10.1002/ajmg.a.32141
- **Therefore, we are unable to assess this variant.**
- **Note:** Since this variant has no confirmed pathomechanism with functional evidence, we cannot consider it for further assessment (we discourage pursuing ASO development without a complete understanding of disease and variant pathomechanisms). However, since exon skipping can be used for both GoF and LoF variants, one can consider it for this variant. The exon is in-frame, does not

code for a stop codon when skipped, and codes for 2.5% of the coding transcript. However, this exon does code for a functionally proven important domain (DOI: 10.1126/science.aau2486). Furthermore, multiple missense variants affecting this domain are thought to be gain-of-function. This suggests an importance for the domain this exon encodes for. Theoretically, it is possible to consider skipping this exon containing the GoF variant in an allele-specific manner, but we would suggest this exon be classified as non-eligible for canonical exon skipping.

**Variant 12: *KIF1A* - NM_001244008.2:c.914C>T**

- Variant description is correct.
- ASO exists, and has been used for a patient: https://www.nlorem.org/patients/gene-programs/
- **Therefore, this variant is considered eligible for knockdown.**

**Variant 13:*CFTR* - NM_000492.4:c.2989-313A>T**

- Variant description is correct, no ASO exists.
- Variant is inherited in an autosomal recessive manner.
- Variant is deep intronic and creates a 118 base pair pseudoexon.
- **Variant is therefore likely eligible for splice correcting ASOs.**

**Variant 14: *HCN2* - NM_001194.4:c.736G>A**

- Variant description is correct, no ASO exists.
- Variant is inherited in an autosomal dominant manner.
- Variant is gain-of-function: https://pubmed.ncbi.nlm.nih.gov/29064616/
- Haploinsufficiency is not a known cause of disease. Loss-of-function missense variant has been reported (https://onlinelibrary.wiley.com/doi/10.1111/epi.17777), but a complete loss of a gene copy is not thought to cause disease.
- **Variant is therefore likely eligible towards knockdown ASOs.**
- **Note:** This variant has been classified as a risk allele in ClinVar. While we'd argue that this classification does not stand given the published literature (https://pubmed.ncbi.nlm.nih.gov/38562733/ and https://pubmed.ncbi.nlm.nih.gov/29064616/), those who would classify the variant as unable to assess on the account of the variant not being pathogenic/likely pathogenic would also be correct.

**Variant 15: *GLDC* - NM_000170.3:c.538C>T**

- Variant description is correct, no ASO exists.
- Variant is inherited in an autosomal recessive manner and the variant is a loss-of-function variant.
- Variant can be assessed for canonical exon skipping.
- Variant located in exon 4. Exon 4 is in-frame and encodes less than 10% of the coding transcripts, however, skipping of exon 4 results in the formation of a premature stop codon (TGA).
- **Therefore, this variant is not eligible for canonical exon skipping.**
